# Multimodal Characterization of High-risk PH-HFpEF phenogroup with Right Ventricular Dysfunction: Vascular Mechanics and Myocardial Transcriptomics

**DOI:** 10.1101/2025.05.15.25327467

**Authors:** Farhan Raza, Zachery R. Gregorich, Jack Freeman, Timothy Houston, Mariana Garcia-Arango, Christopher Lechuga, Yimin Chen, Aditya Sahai, Ahmed El Shaer, Claudia Korcarz, Kai Cui, Yeonhee Park, Kathryn Jones, Wanxin Tu, James Runo, Jefree J. Schulte, Prashant Nagpal, Ying Ge, Naomi Chesler, Ron Stewart, Oliver Wieben, Wei Guo

**Author notes:** To whom correspondence should be addressed: Dr. Farhan Raza, Clinical Science Center, 600 Highland Ave., Madison, WI 53792, United States., And Dr. Wei Guo, 1933 Observatory Drive, 2112 Meat Science & Animal Biologics Discovery Building, Madison, WI 53706, United States. Combined first authors (Farhan Raza, Zachery R. Gregorich).

## Abstract

**Background:** Pulmonary hypertension due to heart failure with preserved ejection fraction (PH-HFpEF) is a highly heterogeneous disease associated with right ventricular (RV) failure and adverse outcomes. Current diagnostic tools inadequately characterize pulmonary vascular disease and RV dysfunction, limiting treatment precision. We hypothesized that deep phenotyping—including invasive hemodynamics, advanced imaging, and myocardial transcriptomics—would identify high-risk PH-HFpEF phenotypes with distinct clinical, physiologic, and molecular characteristics.

**Methods:** 42 PH-HFpEF participants (and 25 pre-capillary PH participants, as a comparison group) underwent clinical evaluation, echocardiography, cardiac MRI (cMRI), and invasive cardiopulmonary exercise testing (iCPET). K-means clustering considering clinical, iCPET, and cMRI data stratified participants into distinct clusters (phenogroups). A subset underwent further characterization by invasive pulmonary vascular mechanics (n=17 PH-HFpEF, n=5 pre-capillary PH) and 4D flow cMRI (n=10 PH-HFpEF, n=5 pre-capillary PH). Endomyocardial biopsies from 10 PH-HFpEF participants were analyzed using long-read RNA-sequencing for differential gene and transcript expression.

**Results:** Clustering revealed two PH-HFpEF phenogroups with significantly different outcomes at one-year follow-up (HR=11.96, CI: 2.66–53.86). High-risk participants had greater left ventricular mass, reduced RV ejection fraction, worse exercise-induced PH, impaired gas exchange (↓peak oxygen consumption, ↑slope of minute ventilation/carbon dioxide production), and lower myocardial strain. Pulmonary vascular mechanics showed higher proximal pulmonary artery stiffness (↑characteristic impedance), increased RV energy expenditure (↑compression waves), and abnormal distal vascular reflections (↓diastolic reflection index) in the high-risk group. 4D flow MRI revealed disturbed flow in pulmonary vasculature (both arteries and veins) in high-risk participants. Transcriptomic analysis implicated differences in post-transcriptional RNA processing and translational control, as well as mitochondrial function, in phenotypic divergence between these HFpEF subgroups. In particular, differential transcript usage for the *TTN* gene, encoding the giant sarcomeric protein titin, may be particularly relevant to elevated myocardial stiffness, which worsens vascular remodeling and precipitates RV dysfunction and failure.

**Conclusions:** In PH-HFpEF, unsupervised clustering using deep physiologic and imaging data identified a high-risk group with impaired RV function and unique transcriptomic profiles. Our findings suggest that proximal and distal pulmonary vascular remodeling, as well as differences in titin isoform expression, may underlie RV failure in this phenogroup. This comprehensive approach provides a framework for mechanistically driven precision medicine strategies in PH-HFpEF.

**Clinical perspectives:** *What is new?:*

- Unsupervised clustering integrating exercise hemodynamics, cardiac MRI, and clinical features revealed a distinct PH-HFpEF subgroup at significantly higher risk for adverse outcomes (HR = 11.96), independent of traditional IpcPH/CpcPH classification.
- Invasive wave mechanics (impedance and wave separation analyses) and non-invasive 4D flow cardiopulmonary MRI reveal unique patterns of proximal stiffness, RV energy inefficiency, and abnormal distal reflections in high-risk PH-HFpEF, providing insights into segmental vascular dysfunction.
- Long-read RNA sequencing of endomyocardial biopsies from PH-HFpEF patients revealed distinct transcriptomic signatures between high- and low-risk PH-HFpEF phenogroups.
- Long-read RNA-seq identified distinct transcript usage of gene TTN, encoding a giant sarcomeric protein (titin) that is a key determinant for myocardial stiffness, suggesting a potential underlying mechanism for RV dysfunction in PH-HFpEF.

*What are clinical implications?:*

- These findings offer a framework for more refined patient classification beyond traditional PVR-based methods, which may improve identification of high-risk individuals who require closer monitoring or targeted therapy.
- The integrative approach highlights titin isoform imbalance and vessel-specific stiffness as potential therapeutic targets, supporting the development of biology-informed, phenotype-specific interventions for PH-HFpEF with RV dysfunction.

## 1. Introduction

Heart failure with preserved ejection fraction (HFpEF) is a complex clinical syndrome characterized by left ventricular diastolic dysfunction and increased myocardial stiffness.^1–4^ HFpEF is rapidly becoming the predominant form of heart failure and its prevalence continues to rise due to the aging population and increasing rates of age-associated comorbidities (e.g., atrial fibrillation, hypertension, diabetes, and obesity).^5–9^ In addition to biventricular myocardial dysfunction,^10^ remodeling of the pulmonary vasculature precipitates pulmonary hypertension (PH) in >83% of HFpEF cases.^11–18^ Pulmonary vascular disease (PVD) in PH-HFpEF eventually leads to right ventricular (RV) failure, worsening symptom burden, exercise intolerance, and mortality.^19–23^ Prevention and management of RV failure remain huge unmet needs as PVD in PH-HFpEF exacerbates heterogeneity in an already complex multi-system disease.^24–27^

The pathobiological nature of PVD in PH-HFpEF is poorly characterized by clinical right heart catheterization, which only captures the steady component of RV afterload with pulmonary vascular resistance (PVR).^21,28–31^ PVR is commonly used to categorize PH-HFpEF into isolated post-capillary PH (IpcPH, with PVR ≤2 WU: Woods unit) or combined pre-/post-capillary PH (CpcPH with PVR >2 WU).^32^ While these categories identify a higher risk PH-HFpEF phenotype (CpcPH),^33,34^ PVR remains a simplistic measure that does not inform vessel-specific pulmonary vascular remodeling.^21,35–37^ This limitation and inability of PVR to better phenotype the heterogenous disease of PH-HFpEF is reflected in multiple failed clinical trials, where various therapies either lacked efficacy or resulted in harm to patients.^26,38–43^

To develop more effective, individualized therapies for PVD in PH-HFpEF, several critical challenges must first be addressed: 1) poor phenotypic and structural characterization of PH-HFpEF with limited data (rest clinical hemodynamics [e.g., PVR] and echocardiogram),^7,21,44^ 2) anatomical heterogeneity of PVD (large pulmonary arteries, small arterioles and capillaries, venules/veins),^21,28,44–46^ and 3) unknown biological mechanisms associated with the transition from low-risk (no RV failure) to a high-risk (with RV failure) PH-HFpEF state.^2,47,48^ Bridging these knowledge gaps is essential for advancing personalized medicine and improving clinical outcomes in patients with PH-HFpEF.

In this study, we implemented a stepwise, multimodal approach to address these gaps. First, we leveraged rich datasets to classify individuals with PH-HFpEF into high-risk vs low-risk groups with invasive exercise hemodynamics and cardiac MRI (cMRI) data. Second, we provided a framework for in-depth pulmonary vessel- and cardiac chamber-specific abnormalities (vascular mechanics flow dynamics and morphologic MRI models) among high-risk vs low-risk PH-HFpEF phenogroups. Lastly, we employed third generation long-read RNA-sequencing (RNA-seq) to profile gene and transcript expression in myocardial biopsies collected from high-risk and low-risk PH-HFpEF participants to identify phenogroup-specific molecular features. Through deep characterization of abnormal mechanics of ventricles (cardiac morphology), pulmonary vascular remodeling (vessel-specific), and biological mechanisms, this study provides a novel framework to gain deeper insights into the pathophysiology of RV failure and PH in HFpEF and uncover potential therapeutic targets for this complex, heterogenous and challenging patient population.

## 2. Methods

### 2.1 Study design and population

This two-step study included a retrospective cohort of 42 PH-HFpEF participants, and a comparison group of 25 pre-capillary PH participants, undergoing in-depth assessment via PH clinic at the University of Wisconsin-Madison. Assessments included clinical evaluations, echocardiography, cMRI, and invasive cardiopulmonary exercise testing (iCPET). In the overall cohort of participants undergoing iCPET (n=190, including 114 PH-HFpEF and 52 pre-capillary PH), only 67 met inclusion criteria as they had a cMRI within 6-months of the iCPET study. Notably, all participants in the prospective arm of the study (mentioned below) had a cardiac MRI within 48 hours of the iCPET study.

All PH-HFpEF participants met the criteria for HFpEF and secondary PH according to the ESC/ERS 2022 PH guidelines,^32^ with heart failure symptoms and a left ventricular (LV) ejection fraction (EF) ≥50% per echocardiogram. Similarly, all participants in pre-capillary PH group met the ESC/ERS 2022 PH guidelines.^32^ For all participants, adverse clinical outcomes (heart failure hospitalization and death) were recorded for one-year after iCPET study.

The second prospective arm of the study included 17 PH-HFpEF and 5 pre-capillary PH participants (these were included in analyses in first arm of the study mentioned above). These participants in prospective arm of the study underwent invasive ventricular-vascular mechanics studies with exercise, which included simultaneous pulmonary artery (PA) pressure and echocardiographic Doppler acquisition (for impedance and wave mechanics analyses) and RV pressure-volume loops.^21,49,50^ Within this prospective cohort, 10 PH-HFpEF and 5 pre-capillary PH participants underwent 4D flow cMRI. An additional 10 PH-HFpEF participants were enrolled in a sub-study for endomyocardial biopsy.

Inclusion criteria for the study group (PH-HFpEF) were as follows: adult patients (≥18 years of age) with confirmed HFpEF and PH, who were referred to our institution for advanced cardiac evaluation. All participants underwent a cMRI and iCPET. Exclusion criteria included: (1) significant valvular heart disease (>moderate mitral or aortic), or other primary causes of heart failure (amyloidosis, hypertrophic cardiomyopathy, prior heart failure with reduced ejection fraction); (2) chronic lung disease requiring supplemental oxygen (chronic obstructive pulmonary disease, interstitial lung disease); (3) active malignancy or recent cancer treatment; (4) severe renal impairment (eGFR <30 mL/min/1.73 m²); and (5) other conditions that could confound the assessment of cardiac function (e.g., severe anemia, acute decompensated heart failure). All participants provided written informed consent in accordance with University of Wisconsin-Madison institutional review board approval. The study complied with the Declaration of Helsinki.

### 2.2 Invasive Cardiopulmonary Exercise Test

iCPET was performed using a recumbent stationary ergometer, as previously reported.^16–18,21,34,43,50^ Briefly, right heart catheterization was performed with a balloon-tipped catheter through a 7-Fr venous sheath via internal jugular access and a 4-Fr radial arterial catheter was placed for arterial blood gas analyses. The breath-by-breath expired gas exchange analysis was performed with an Ultima™ CardiO2® MedGraphics metabolic cart, which was connected to participants via a mouthpiece and nose-clip to avoid air leak. Cardiac output was measured at all stages with direct Fick principle. Hemodynamic data was acquired at rest, different stages of exercise (every 25 watts, peak-exercise data reported). Specific CPET metrics were recorded: peak oxygen consumption (VO2), slope of minute ventilation/carbon dioxide production (VE/VCO2) and end-tidal carbon dioxide from rest-to-exercise (ETCO2).

### 2.3 Cardiac MRI

cMRI was performed on all participants to assess cardiac structure and function. Imaging was conducted using a 3.0-T MRI scanner (Signa Premier, GE Healthcare, Waukesha, WI) at the University of Wisconsin-Madison. Standard cine imaging sequences were obtained to evaluate LV and RV volumes, EF, and myocardial mass. LV and RV function was specifically assessed using short-axis and axial cine balanced steady-state free precession views to quantify RV volumes and EF, as previously described.^21,51^ Scan parameters included reconstructed temporal resolution = 35-45 ms, repetition time = 3.1 ms, echo time = 1.1 ms, field of view = 40 × 40 cm, acquired spatial resolution = 1.79 × 1.79 mm, reconstructed spatial resolution = 1.56 × 1.56 mm, slice thickness = 8 mm, and reconstructed cardiac phases = 20.

### 2.4 Integrative cardiopulmonary 4D flow MRI

A subset of 15 participants underwent 4D flow cMRI to assess pulmonary vascular and RV flow patterns (10 PH-HFpEF and 5 pre-capillary PH). Imaging was performed using a 3.0 T MRI scanner [GE SIGNA™ Premier], with a radially undersampled 4D flow cMRI protocol.^52^ This technique allows for a comprehensive, retrospective, detailed visualization and quantification of blood flow dynamics in the vasculature and cardiac chambers of the heart. The 4D flow data were repeated after a vasodilator challenge (0.4mg sublingual nitroglycerine) in 6 PH-HFpEF participants.

4D flow cMRI data were pre-processed following established guidelines accounting for noise corrections, background phase corrections, and velocity anti-aliasing.^53^ A 3D PC angiogram was then generated to manually segment the pulmonary veins, pulmonary arteries, left atrium (LA), and aorta using commercial software (Mimics Innovation Suite 20.0, Materialize, Leuven, Belgium). The manual segmentations were then exported to a flow quantification software (EnSight 2022 R2, Ansys, Canonsburg, PA) where cut planes were placed at standardized regions of interest for each vessel (aorta, pulmonary artery and pulmonary veins) and cardiac chambers (left atrium left ventricle), followed by creating flow streamline visualizations. The cut planes were then processed using custom in-house software in MATLAB (Mathworks, Natick, MA).

### 2.5 Impedance and wave intensity analysis (WIA)

For quantification of impedance and WIA, simultaneous pulmonary artery pressure (via Millar catheter in the pulmonary artery during iCPET) and RV outflow tract pulse wave Doppler-based flow (obtained via Doppler echocardiography) were collected at rest and during low-workload exercise (25 watts), as previously described.^21,50^ These simultaneous pressure-flow data were post-processed using a time-domain method to generate a PQ-loop (pressure-flow).^54,55^ This loop was then used to derive metrics that specify vessel-specific and cardiac-cycle-specific abnormalities, as well as ventricular-vascular interactions in the pulmonary circulation and RV.^55,56^ These metrics included characteristic impedance (a measure of proximal pulmonary artery stiffness),^21,31^ compression waves during systole (which reflect RV force generation and proximal wave reflections from the pulmonary circulation), and decompression waves during diastole (representing RV relaxation and distal vascular reflections from pulmonary venular/venous segments).^45,57–64^ We have described these methods previously in detail.^55,56^

### 2.6 Endomyocardial biopsy collection

Endomyocardial septal biopsies were collected from a subset of participants (PH-HFpEF=10) to define differential gene and transcript expression associated with high-risk state of RV failure (vs. low-risk state with preserved RV function) in PH-HFpEF. Briefly, after iCPET, participants recovered to their baseline and then given moderate sedation. The 7Fr short sheath for iCPET was replaced over the wire with a long-curved sheath (7FR x 45cm Argon Axcess^TM^ Introducer sheath: SKU 191745), which was placed under fluoroscopy and balloon-tipped catheter towards the direction of interventricular septum to prevent any complications of RV free wall damage with biopsy. Through this long-curved sheath, a bioptome (6Fr SparrowHawk 1.8mm x 50 cm biopsy forcep) was advanced through the sheath and into the RV under fluoroscopic guidance. The bioptome was used to obtain small tissue samples from the interventricular septal endomyocardium (Figure S1). Multiple tissue samples (typically 4) were obtained and immediately placed in liquid nitrogen. Following biopsy collection, the sheath was removed, and manual compression was applied to the puncture site to ensure hemostasis. The patient was monitored for any complications in the recovery area for one-hour post-procedure.

### 2.7 Long-read RNA-seq

To assess differences in gene and transcript expression between individuals in the low- and high-risk PH-HFpEF phenogroups PacBio long-read RNA-seq was carried out as follows.

Total RNA was extracted and prepared into full-length cDNA libraries using the Kinnex Full-Length RNA Kit, following the manufacturer’s instructions. These libraries were sequenced on a PacBio Revio instrument (Pacific Biosciences) using a 30-hour movie run, yielding long-read RNA-seq data. Primary data processing (including segmentation, demultiplexing, and refinement) was performed on the Revio instrument with SMRT Link v13.1 to produce high-quality full-length reads.

The generated FASTQ files were then analyzed with IsoQuant v3.6.1 using the default gene quantification strategy and the “with ambiguous” transcript quantification strategy. IsoQuant was run using the human genome reference (GRCh38) and annotation (Homo sapiens.GRCh38.113)^65^. Minimap2 v2.28 was employed internally by IsoQuant for read alignment, and subsequent sorting and indexing of BAM files were performed. IsoQuant produced both transcript-level and aggregated gene-level count matrices for each sample^66^.

After generating gene- and transcript-level count matrices, we performed differential expression analysis using the DESeq2 R package (v1.42.1) in R (v4.3.3)^67^. Before analysis, we filtered out genes with a mean raw count below 10 and transcripts with counts below 10 in all samples from either the high-risk or low-risk phenogroup. DESeq2’s standard workflow was applied, including size factor estimation, dispersion estimation, and fitting of a negative binomial model. Outlier detection and replacement were performed with default parameters, followed by final model refitting as needed. Differentially expressed genes and transcripts were identified using a contrast comparing the high-risk phenogroup against the low-risk phenogroup. Principal Component Analysis (PCA) was conducted at the gene level using scikit-learn (v1.3.2) in Python to assess the global variance in transcriptomic profiles^68^. The PCA was performed on log2-transformed median-ratio normalized counts, and the first two principal components were visualized to illustrate differences between high- and low-risk phenogroups. In addition, Gene Set Enrichment Analysis (GSEA) results were generated to facilitate pathway enrichment analysis^69^.

For GSEA, we employed the clusterProfiler package (v4.10.0, installed via bioconda) in R (v4.3.3) to perform enrichment analysis on Gene Ontology (GO) terms across biological processes, molecular functions, and cellular components^70^. The DESeq2 ’stat’ value, representing the Wald test statistic for each gene, was used as the ranking metric, capturing both the magnitude and statistical confidence of differential expression. GO enrichment analysis of novel transcripts was also performed using an over-representation analysis approach implemented in clusterProfiler, considering genes exhibiting greater than 0.1% novel transcript usage in the high- and low-risk phenogroups. Significant GO terms were identified based on an adjusted p-value threshold of 0.05. Results were visualized using bar plots to highlight enriched biological pathways distinctly associated with each phenogroup.

### 2.8 Statistical analysis

Baseline characteristics were presented as mean ± SD for continuous variables and as frequencies and proportions (%) for categorical variables. For continuous variables, a comparison among groups was conducted using the analysis of variance (ANOVA) test (or Wilcoxon rank sum test for non-normal distribution). Differences in categorical data were assessed using the χ2 statistic or Fisher exact test, as appropriate. For clustering analyses, the K-means clustering algorithm was used, which is an unsupervised learning method that partitioned the dataset into K-clusters based on the similarity of observations. The optimal number of clusters was determined using both the elbow method and silhouette analysis, both of which consistently indicated that two clusters provided the best fit to the data, based on proximity to the computed centroids. Clustering analyses were repeated on progressively complex variable sets (Table S1). Initially, clusters were formed using clinical variables. The model was then extended to include both rest and exercise hemodynamics data, then followed by the addition of cardiac MRI data. For data processing prior to clustering, the data were standardized to ensure that all variables were on the same scale. Kaplan-Meier survival curves were subsequently created for the IpcPH/CpcPH cluster and three types of K-means clusters to estimate survival rates within each group. Differences between groups were evaluated using the log-rank test. Additionally, multivariate Cox proportional hazard regression, adjusted for age and sex, was used to identify and quantify the association between predictors and the hazard of an event. The analyses were conducted in R version 4.4.2 and Python version 3.9.7. For the analysis of RNA-seq data via DESeq2, GSEA, and GO enrichment, p-values were adjusted using the Benjamini and Hochberg method.^71^

## 3. Results

### 3.1 K-means clustering for deep phenotyping and defining high-risk phenogroups in PH-HFpEF

To define distinct PH-HFpEF phenogroups using a rich dataset, all participants (n=42) were classified into two distinct phenogroups using a K-means clustering approach, based on progressively complex variable sets (Figure 1 and Table S1). In the first clustering analysis, using only standard clinical variables, two unique clusters were identified that demonstrated significantly different clinical outcomes (Hazard ratio=2.90, CI: 1.28-6.56). This stratification was comparable to invasive right heart catheterization-guided current standard of PVR-based PH-HFpEF grouping into IpcPH vs CpcPH (Hazard ratio=2.67, CI: 1.02-7.01). The second clustering analysis, which incorporated the iCPET data to clinical variables, generated two clusters with further stratification and more divergent clinical outcomes (Hazard ratio=4.31, CI: 1.88-9.91). In the third clustering analysis by adding cMRI data (to clinical and iCPET data) produced two phenogroups with the most pronounced difference in clinical outcomes (Hazard ratio=11.96, CI: 2.66-53.86), underscoring the additive prognostic value of integrated multimodal phenotyping in PH-HFpEF.

**Figure 1.**
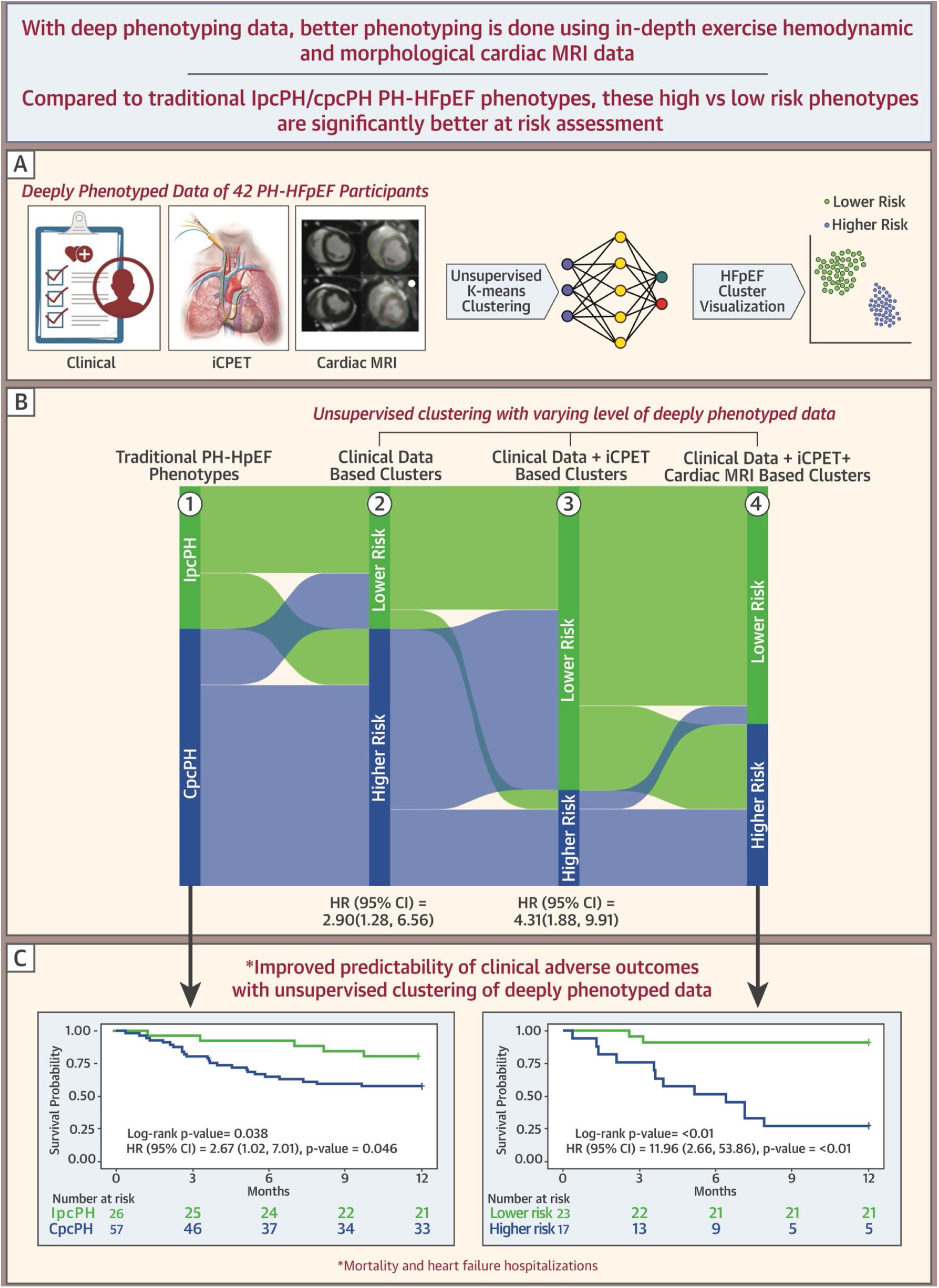
K-means clustering to define high- vs low-risk PH-HFpEF phenogroups and Sankey diagram to represent redistribution.

### 3.2 Features of low- vs. high-risk PH-HFpEF phenogroups

Based on the stepwise clustering approach above, two distinct phenogroups were defined and their characteristics are summarized in Table 1. There was a trend (statistically non-significant in most cases) in high-risk PH-HFpEF participants to be older, male, with a lower BMI and higher incidence of atrial fibrillation. On echocardiograms, these individuals had increased LV mass, LA size, RV:PA uncoupling, RV dilation, and dysfunction. Hemodynamics revealed lower cardiac index and pulmonary arterial compliance at rest and with exercise, and an increased mean pulmonary arterial pressure/cardiac output (mPAP/CO) slope (worse exercise PH). Cardiopulmonary exercise data revealed decreased aerobic capacity (lower VO2, oxygen uptake efficiency slope [OUES]) and poor ventilatory efficiency (VE/VCO2). cMRI revealed an increased RV size, lower RVEF, decreased biventricular strain, and increased septal angle (signifying increased interventricular dependence).

**Table 1.**
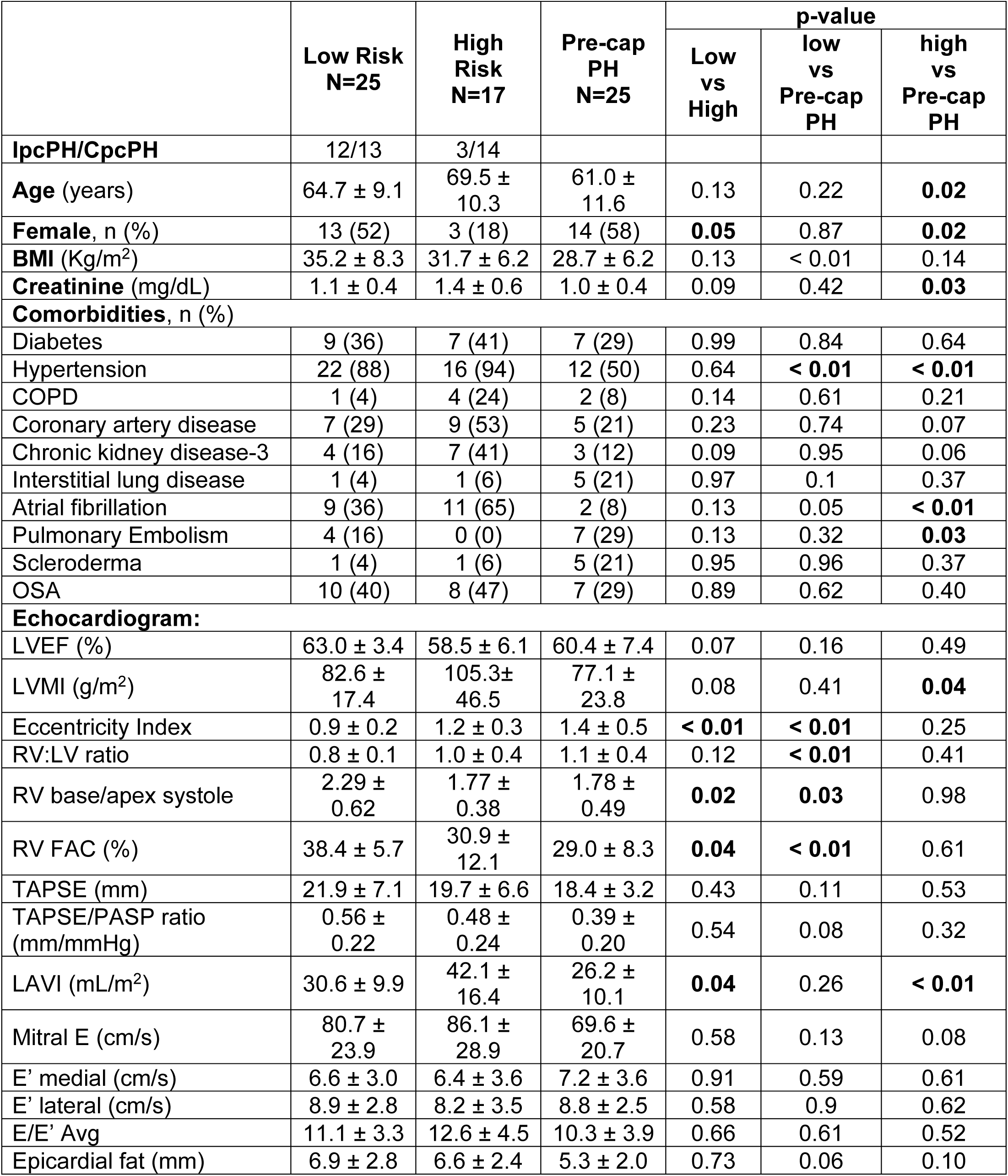
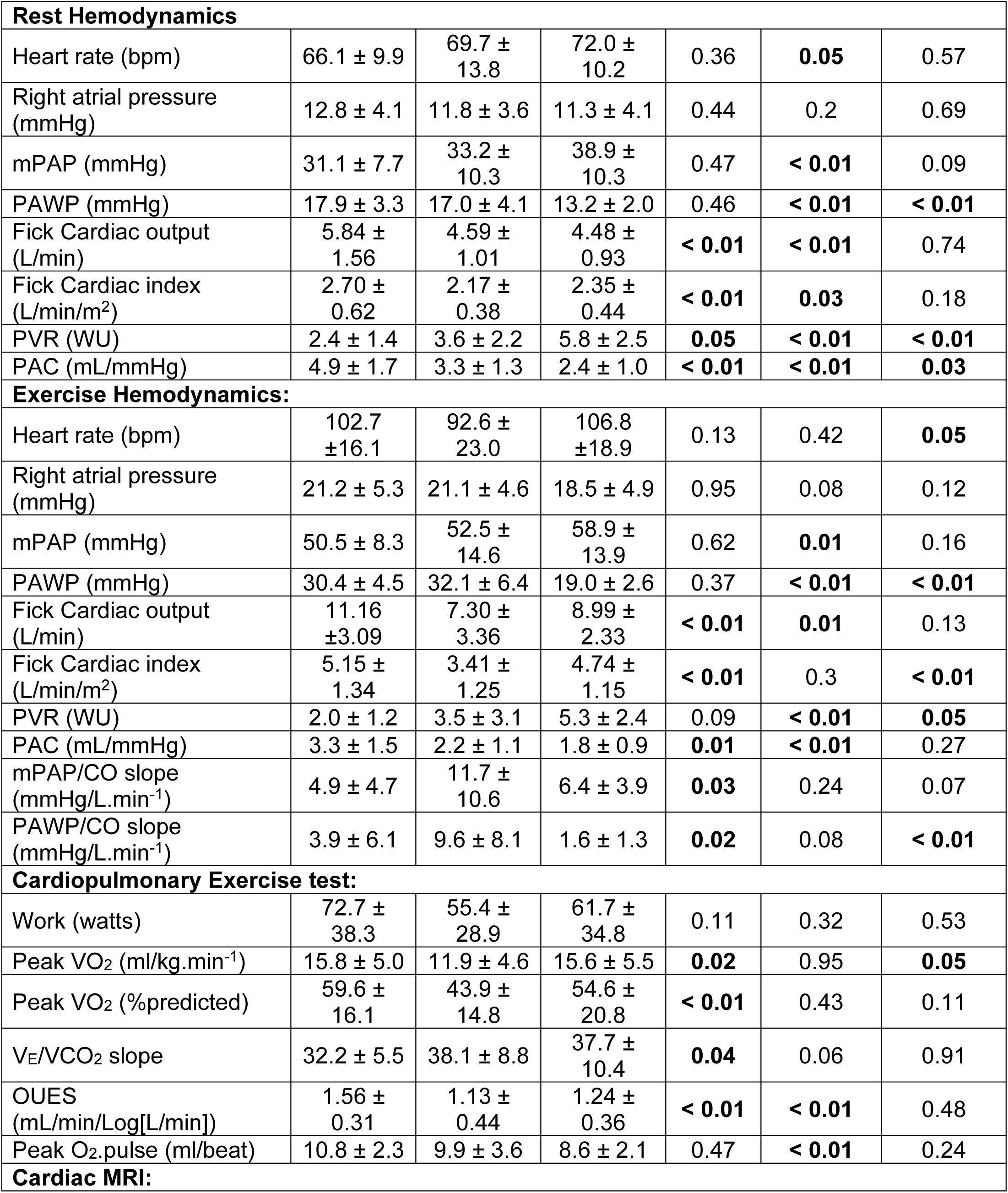
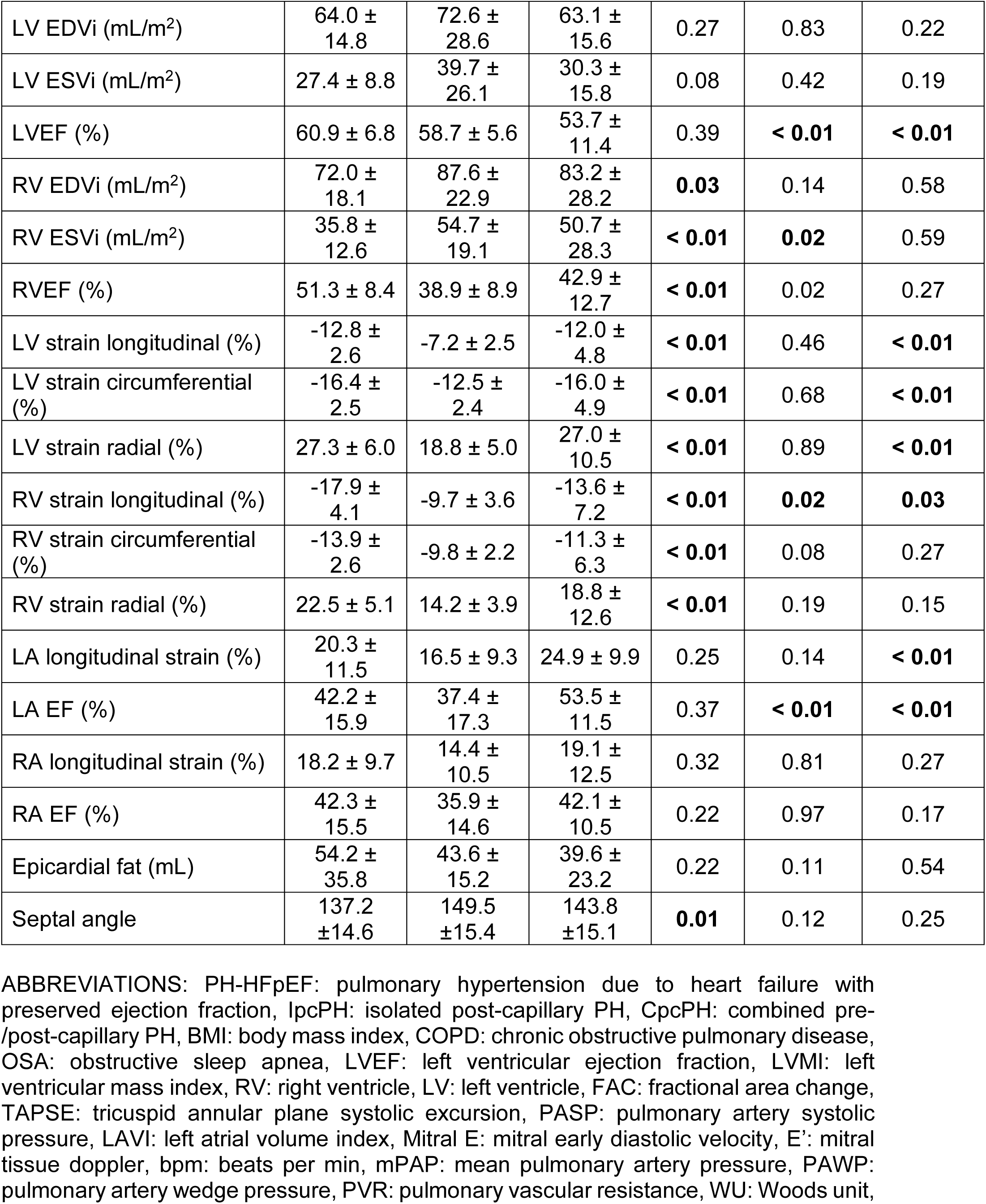
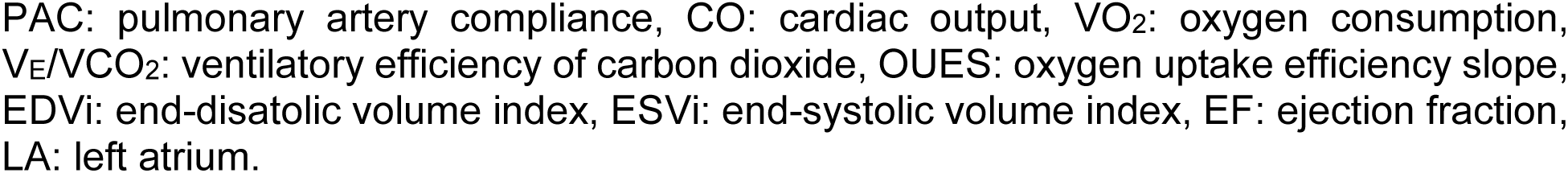
Deeply phenotyped PH-HFpEF phenogroups and comparison pre-capillary PH group.

### 3.3 Features of re-classified traditional IpcPH/CpcPH groups into high- and low-risk PH-HFpEF phenogroups

While there was an overlap of conventional PH phenotypes (IpcPH/CpcPH) among the low- vs. high-risk PH-HFpEF phenogroups, there was significant redistribution of traditional PH phenotypes into different risk phenogroups (Figure 1). To better understand this redistribution, subgroup comparison is summarized in Table 2.

**Table 2.**
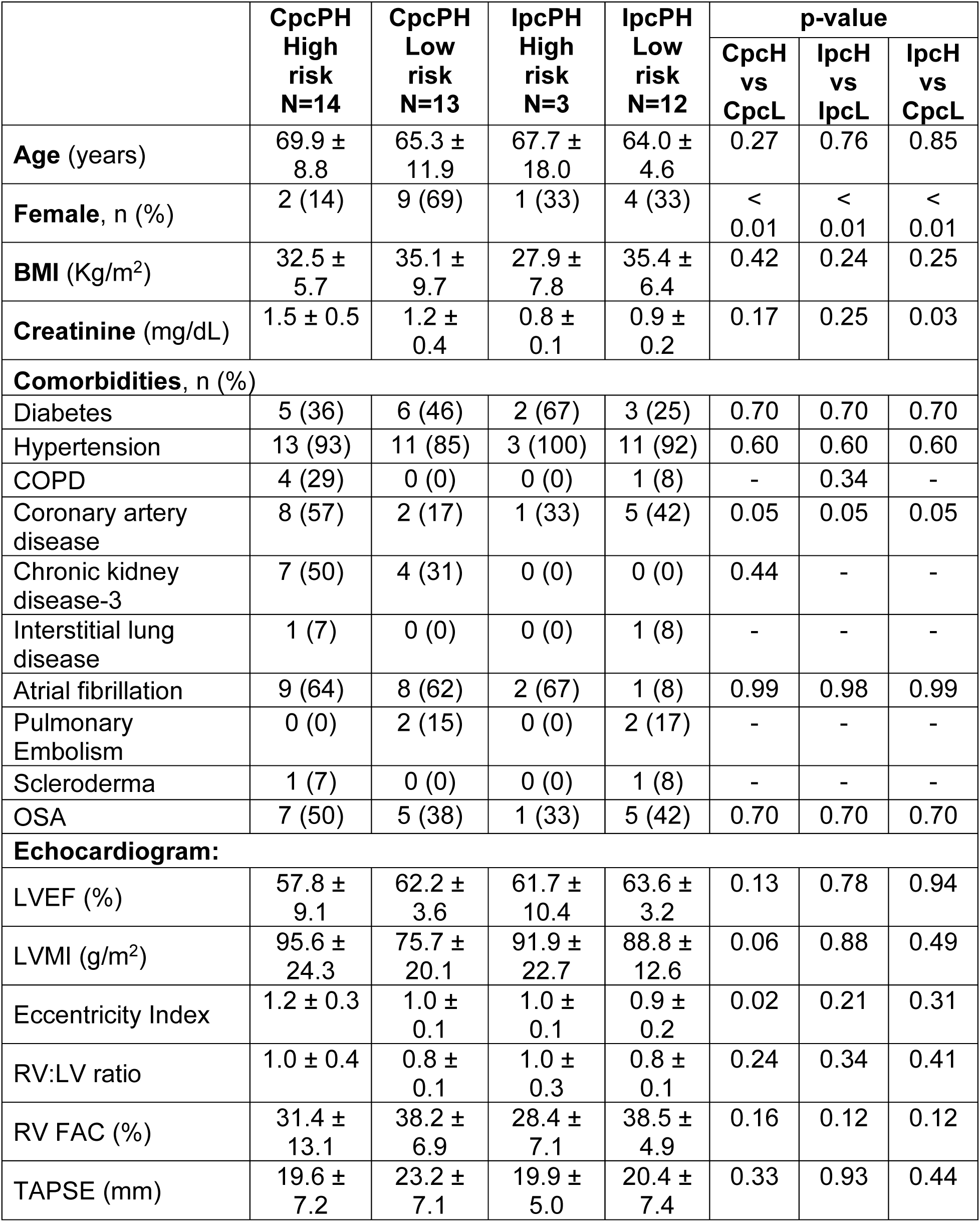
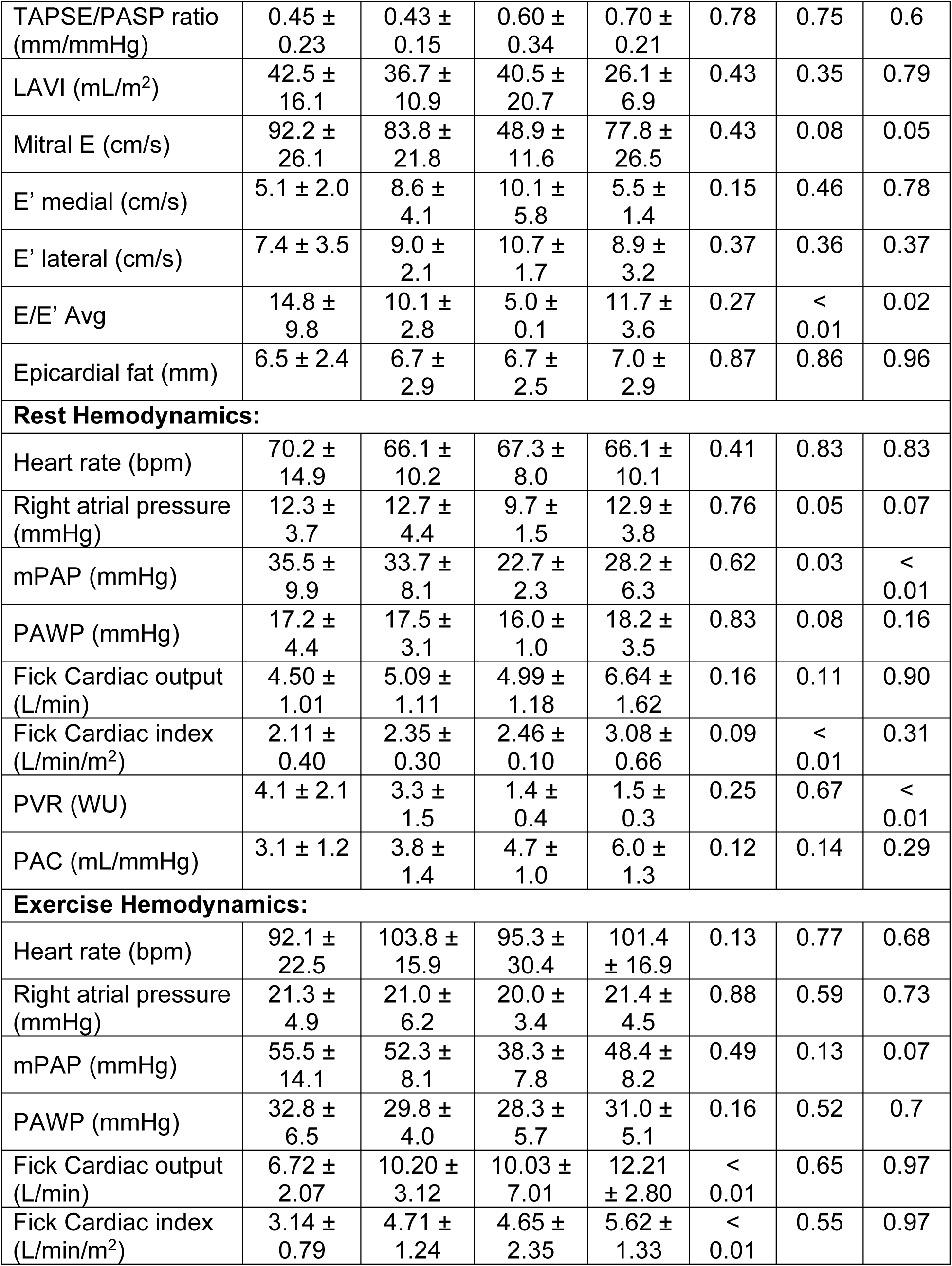
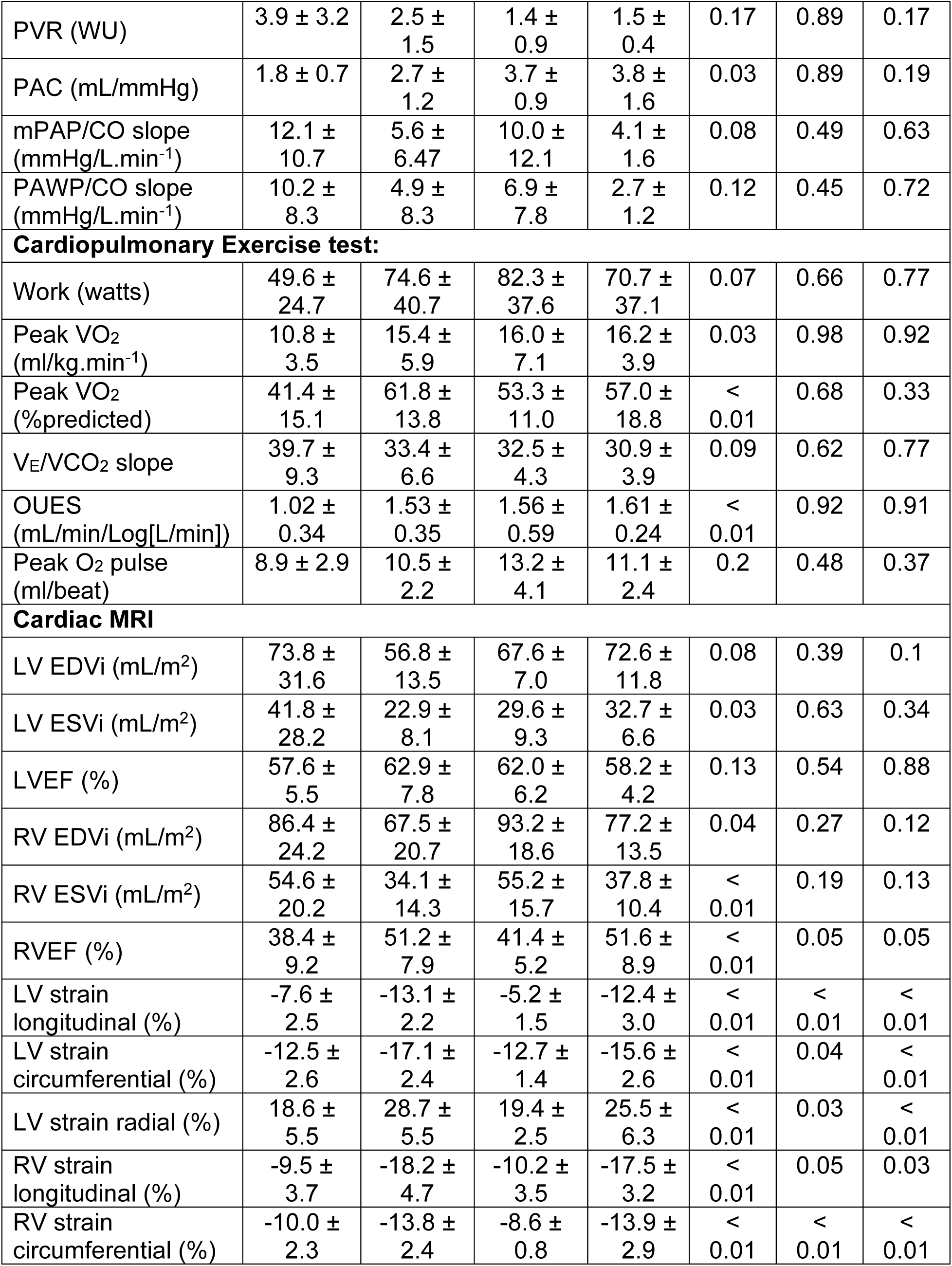
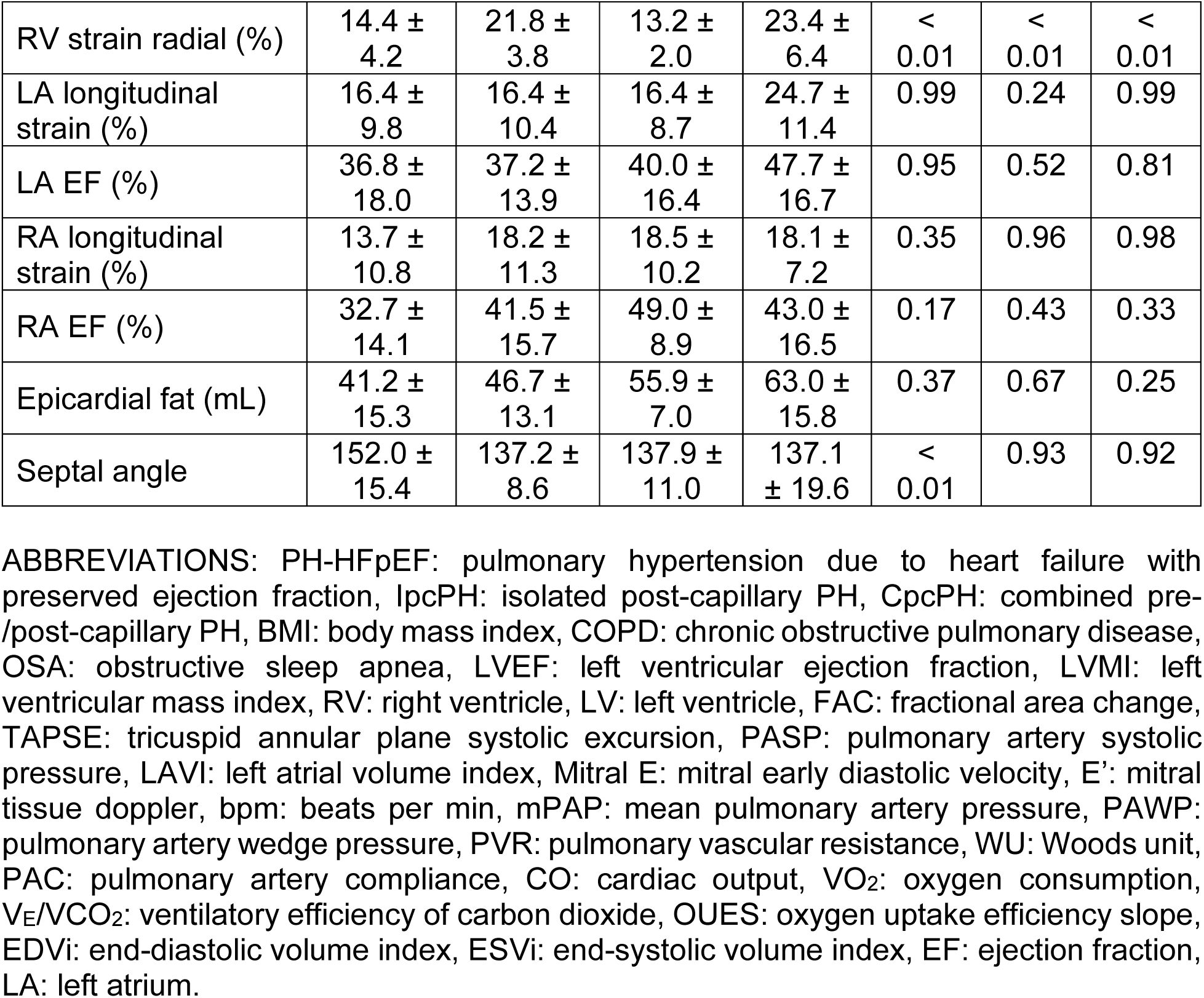
Reclassification of conventional Ipc vs CpcPH patients into appropriate risk category. Traditional IpcPH/CpcPH phenotypes are reclassified into different high vs low risk categories based on deep phenotyping and unsupervised clustering algorithm.

|  | **CpcPH**<br>**High**<br>**risk**<br>**N=14** | **CpcPH**<br>**Low**<br>**risk**<br>**N=13** | **lpcPH**<br>**High**<br>**risk**<br>**N=3** | **lpcPH**<br>**Low**<br>**risk**<br>**N=12** | **p-value** |  |  |
| --- | --- | --- | --- | --- | --- | --- | --- |
|  |  |  |  |  | **CpcH**<br>**vs**<br>**CpcL** | **lpcH**<br>**vs**<br>**lpcL** | **lpcH**<br>**vs**<br>**CpcL** |
| **Age (years)** | 69.9 ± 8.8 | 65.3 ± 11.9 | 67.7 ± 18.0 | 64.0 ± 4.6 | 0.27 | 0.76 | 0.85 |
| **Female, n (%)** | 2 (14) | 9 (69) | 1 (33) | 4 (33) | < 0.01 | < 0.01 | < 0.01 |
| **BMI (Kg/m<sup>2</sup>)** | 32.5 ± 5.7 | 35.1 ± 9.7 | 27.9 ± 7.8 | 35.4 ± 6.4 | 0.42 | 0.24 | 0.25 |
| **Creatinine (mg/dL)** | 1.5 ± 0.5 | 1.2 ± 0.4 | 0.8 ± 0.1 | 0.9 ± 0.2 | 0.17 | 0.25 | 0.03 |
| **Comorbidities, n (%)** |  |  |  |  |  |  |  |
| Diabetes | 5 (36) | 6 (46) | 2 (67) | 3 (25) | 0.70 | 0.70 | 0.70 |
| Hypertension | 13 (93) | 11 (85) | 3 (100) | 11 (92) | 0.60 | 0.60 | 0.60 |
| COPD | 4 (29) | 0 (0) | 0 (0) | 1 (8) | - | 0.34 | - |
| Coronary artery disease | 8 (57) | 2 (17) | 1 (33) | 5 (42) | 0.05 | 0.05 | 0.05 |
| Chronic kidney disease-3 | 7 (50) | 4 (31) | 0 (0) | 0 (0) | 0.44 | - | - |
| Interstitial lung disease | 1 (7) | 0 (0) | 0 (0) | 1 (8) | - | - | - |
| Atrial fibrillation | 9 (64) | 8 (62) | 2 (67) | 1 (8) | 0.99 | 0.98 | 0.99 |
| Pulmonary Embolism | 0 (0) | 2 (15) | 0 (0) | 2 (17) | - | - | - |
| Scleroderma | 1 (7) | 0 (0) | 0 (0) | 1 (8) | - | - | - |
| OSA | 7 (50) | 5 (38) | 1 (33) | 5 (42) | 0.70 | 0.70 | 0.70 |
| **Echocardiogram:** |  |  |  |  |  |  |  |
| LVEF (%) | 57.8 ± 9.1 | 62.2 ± 3.6 | 61.7 ± 10.4 | 63.6 ± 3.2 | 0.13 | 0.78 | 0.94 |
| LVMI (g/m<sup>2</sup>) | 95.6 ± 24.3 | 75.7 ± 20.1 | 91.9 ± 22.7 | 88.8 ± 12.6 | 0.06 | 0.88 | 0.49 |
| Eccentricity Index | 1.2 ± 0.3 | 1.0 ± 0.1 | 1.0 ± 0.1 | 0.9 ± 0.2 | 0.02 | 0.21 | 0.31 |
| RV:LV ratio | 1.0 ± 0.4 | 0.8 ± 0.1 | 1.0 ± 0.3 | 0.8 ± 0.1 | 0.24 | 0.34 | 0.41 |
| RV FAC (%) | 31.4 ± 13.1 | 38.2 ± 6.9 | 28.4 ± 7.1 | 38.5 ± 4.9 | 0.16 | 0.12 | 0.12 |
| TAPSE (mm) | 19.6 ± 7.2 | 23.2 ± 7.1 | 19.9 ± 5.0 | 20.4 ± 7.4 | 0.33 | 0.93 | 0.44 |
| TAPSE/PASP ratio (mm/mmHg) | 0.45 ± 0.23 | 0.43 ± 0.15 | 0.60 ± 0.34 | 0.70 ± 0.21 | 0.78 | 0.75 | 0.6 |
| LAVI (mL/m<sup>2</sup>) | 42.5 ± 16.1 | 36.7 ± 10.9 | 40.5 ± 20.7 | 26.1 ± 6.9 | 0.43 | 0.35 | 0.79 |
| Mitral E (cm/s) | 92.2 ± 26.1 | 83.8 ± 21.8 | 48.9 ± 11.6 | 77.8 ± 26.5 | 0.43 | 0.08 | 0.05 |
| E' medial (cm/s) | 5.1 ± 2.0 | 8.6 ± 4.1 | 10.1 ± 5.8 | 5.5 ± 1.4 | 0.15 | 0.46 | 0.78 |
| E' lateral (cm/s) | 7.4 ± 3.5 | 9.0 ± 2.1 | 10.7 ± 1.7 | 8.9 ± 3.2 | 0.37 | 0.36 | 0.37 |
| E/E' Avg | 14.8 ± 9.8 | 10.1 ± 2.8 | 5.0 ± 0.1 | 11.7 ± 3.6 | 0.27 | < 0.01 | 0.02 |
| Epicardial fat (mm) | 6.5 ± 2.4 | 6.7 ± 2.9 | 6.7 ± 2.5 | 7.0 ± 2.9 | 0.87 | 0.86 | 0.96 |
| **Rest Hemodynamics:** |  |  |  |  |  |  |  |
| Heart rate (bpm) | 70.2 ± 14.9 | 66.1 ± 10.2 | 67.3 ± 8.0 | 66.1 ± 10.1 | 0.41 | 0.83 | 0.83 |
| Right atrial pressure (mmHg) | 12.3 ± 3.7 | 12.7 ± 4.4 | 9.7 ± 1.5 | 12.9 ± 3.8 | 0.76 | 0.05 | 0.07 |
| mPAP (mmHg) | 35.5 ± 9.9 | 33.7 ± 8.1 | 22.7 ± 2.3 | 28.2 ± 6.3 | 0.62 | 0.03 | < 0.01 |
| PAWP (mmHg) | 17.2 ± 4.4 | 17.5 ± 3.1 | 16.0 ± 1.0 | 18.2 ± 3.5 | 0.83 | 0.08 | 0.16 |
| Fick Cardiac output (L/min) | 4.50 ± 1.01 | 5.09 ± 1.11 | 4.99 ± 1.18 | 6.64 ± 1.62 | 0.16 | 0.11 | 0.90 |
| Fick Cardiac index (L/min/m<sup>2</sup>) | 2.11 ± 0.40 | 2.35 ± 0.30 | 2.46 ± 0.10 | 3.08 ± 0.66 | 0.09 | < 0.01 | 0.31 |
| PVR (WU) | 4.1 ± 2.1 | 3.3 ± 1.5 | 1.4 ± 0.4 | 1.5 ± 0.3 | 0.25 | 0.67 | < 0.01 |
| PAC (mL/mmHg) | 3.1 ± 1.2 | 3.8 ± 1.4 | 4.7 ± 1.0 | 6.0 ± 1.3 | 0.12 | 0.14 | 0.29 |
| **Exercise Hemodynamics:** |  |  |  |  |  |  |  |
| Heart rate (bpm) | 92.1 ± 22.5 | 103.8 ± 15.9 | 95.3 ± 30.4 | 101.4 ± 16.9 | 0.13 | 0.77 | 0.68 |
| Right atrial pressure (mmHg) | 21.3 ± 4.9 | 21.0 ± 6.2 | 20.0 ± 3.4 | 21.4 ± 4.5 | 0.88 | 0.59 | 0.73 |
| mPAP (mmHg) | 55.5 ± 14.1 | 52.3 ± 8.1 | 38.3 ± 7.8 | 48.4 ± 8.2 | 0.49 | 0.13 | 0.07 |
| PAWP (mmHg) | 32.8 ± 6.5 | 29.8 ± 4.0 | 28.3 ± 5.7 | 31.0 ± 5.1 | 0.16 | 0.52 | 0.7 |
| Fick Cardiac output (L/min) | 6.72 ± 2.07 | 10.20 ± 3.12 | 10.03 ± 7.01 | 12.21 ± 2.80 | < 0.01 | 0.65 | 0.97 |
| Fick Cardiac index (L/min/m<sup>2</sup>) | 3.14 ± 0.79 | 4.71 ± 1.24 | 4.65 ± 2.35 | 5.62 ± 1.33 | < 0.01 | 0.55 | 0.97 |
| PVR (WU) | 3.9 ± 3.2 | 2.5 ± 1.5 | 1.4 ± 0.9 | 1.5 ± 0.4 | 0.17 | 0.89 | 0.17 |
| PAC (mL/mmHg) | 1.8 ± 0.7 | 2.7 ± 1.2 | 3.7 ± 0.9 | 3.8 ± 1.6 | 0.03 | 0.89 | 0.19 |
| mPAP/CO slope (mmHg/L.min<sup>-1</sup>) | 12.1 ± 10.7 | 5.6 ± 6.47 | 10.0 ± 12.1 | 4.1 ± 1.6 | 0.08 | 0.49 | 0.63 |
| PAWP/CO slope (mmHg/L.min<sup>-1</sup>) | 10.2 ± 8.3 | 4.9 ± 8.3 | 6.9 ± 7.8 | 2.7 ± 1.2 | 0.12 | 0.45 | 0.72 |
| **Cardiopulmonary Exercise test:** |  |  |  |  |  |  |  |
| Work (watts) | 49.6 ± 24.7 | 74.6 ± 40.7 | 82.3 ± 37.6 | 70.7 ± 37.1 | 0.07 | 0.66 | 0.77 |
| Peak VO<sub>2</sub> (ml/kg.min<sup>-1</sup>) | 10.8 ± 3.5 | 15.4 ± 5.9 | 16.0 ± 7.1 | 16.2 ± 3.9 | 0.03 | 0.98 | 0.92 |
| Peak VO<sub>2</sub> (%predicted) | 41.4 ± 15.1 | 61.8 ± 13.8 | 53.3 ± 11.0 | 57.0 ± 18.8 | < 0.01 | 0.68 | 0.33 |
| V<sub>E</sub>/VCO<sub>2</sub> slope | 39.7 ± 9.3 | 33.4 ± 6.6 | 32.5 ± 4.3 | 30.9 ± 3.9 | 0.09 | 0.62 | 0.77 |
| OUES (mL/min/Log[L/min]) | 1.02 ± 0.34 | 1.53 ± 0.35 | 1.56 ± 0.59 | 1.61 ± 0.24 | < 0.01 | 0.92 | 0.91 |
| Peak O<sub>2</sub> pulse (ml/beat) | 8.9 ± 2.9 | 10.5 ± 2.2 | 13.2 ± 4.1 | 11.1 ± 2.4 | 0.2 | 0.48 | 0.37 |
| **Cardiac MRI** |  |  |  |  |  |  |  |
| LV EDVi (mL/m<sup>2</sup>) | 73.8 ± 31.6 | 56.8 ± 13.5 | 67.6 ± 7.0 | 72.6 ± 11.8 | 0.08 | 0.39 | 0.1 |
| LV ESVi (mL/m<sup>2</sup>) | 41.8 ± 28.2 | 22.9 ± 8.1 | 29.6 ± 9.3 | 32.7 ± 6.6 | 0.03 | 0.63 | 0.34 |
| LVEF (%) | 57.6 ± 5.5 | 62.9 ± 7.8 | 62.0 ± 6.2 | 58.2 ± 4.2 | 0.13 | 0.54 | 0.88 |
| RV EDVi (mL/m<sup>2</sup>) | 86.4 ± 24.2 | 67.5 ± 20.7 | 93.2 ± 18.6 | 77.2 ± 13.5 | 0.04 | 0.27 | 0.12 |
| RV ESVi (mL/m<sup>2</sup>) | 54.6 ± 20.2 | 34.1 ± 14.3 | 55.2 ± 15.7 | 37.8 ± 10.4 | < 0.01 | 0.19 | 0.13 |
| RVEF (%) | 38.4 ± 9.2 | 51.2 ± 7.9 | 41.4 ± 5.2 | 51.6 ± 8.9 | < 0.01 | 0.05 | 0.05 |
| LV strain longitudinal (%) | -7.6 ± 2.5 | -13.1 ± 2.2 | -5.2 ± 1.5 | -12.4 ± 3.0 | < 0.01 | < 0.01 | < 0.01 |
| LV strain circumferential (%) | -12.5 ± 2.6 | -17.1 ± 2.4 | -12.7 ± 1.4 | -15.6 ± 2.6 | < 0.01 | 0.04 | < 0.01 |
| LV strain radial (%) | 18.6 ± 5.5 | 28.7 ± 5.5 | 19.4 ± 2.5 | 25.5 ± 6.3 | < 0.01 | 0.03 | < 0.01 |
| RV strain longitudinal (%) | -9.5 ± 3.7 | -18.2 ± 4.7 | -10.2 ± 3.5 | -17.5 ± 3.2 | < 0.01 | 0.05 | 0.03 |
| RV strain circumferential (%) | -10.0 ± 2.3 | -13.8 ± 2.4 | -8.6 ± 0.8 | -13.9 ± 2.9 | < 0.01 | < 0.01 | < 0.01 |
| RV strain radial (%) | 14.4 ± 4.2 | 21.8 ± 3.8 | 13.2 ± 2.0 | 23.4 ± 6.4 | < 0.01 | < 0.01 | < 0.01 |
| LA longitudinal strain (%) | 16.4 ± 9.8 | 16.4 ± 10.4 | 16.4 ± 8.7 | 24.7 ± 11.4 | 0.99 | 0.24 | 0.99 |
| LA EF (%) | 36.8 ± 18.0 | 37.2 ± 13.9 | 40.0 ± 16.4 | 47.7 ± 16.7 | 0.95 | 0.52 | 0.81 |
| RA longitudinal strain (%) | 13.7 ± 10.8 | 18.2 ± 11.3 | 18.5 ± 10.2 | 18.1 ± 7.2 | 0.35 | 0.96 | 0.98 |
| RA EF (%) | 32.7 ± 14.1 | 41.5 ± 15.7 | 49.0 ± 8.9 | 43.0 ± 16.5 | 0.17 | 0.43 | 0.33 |
| Epicardial fat (mL) | 41.2 ± 15.3 | 46.7 ± 13.1 | 55.9 ± 7.0 | 63.0 ± 15.8 | 0.37 | 0.67 | 0.25 |
| Septal angle | 152.0 ± 15.4 | 137.2 ± 8.6 | 137.9 ± 11.0 | 137.1 ± 19.6 | < 0.01 | 0.93 | 0.92 |

ABBREVIATIONS: PH-HFpEF: pulmonary hypertension due to heart failure with preserved ejection fraction, lpcPH: isolated post-capillary PH, CpcPH: combined pre-/post-capillary PH, BMI: body mass index, COPD: chronic obstructive pulmonary disease, OSA: obstructive sleep apnea, LVEF: left ventricular ejection fraction, LVMI: left ventricular mass index, RV: right ventricle, LV: left ventricle, FAC: fractional area change, TAPSE: tricuspid annular plane systolic excursion, PASP: pulmonary artery systolic pressure, LAVI: left atrial volume index, Mitral E: mitral early diastolic velocity, E': mitral tissue doppler, bpm: beats per min, mPAP: mean pulmonary artery pressure, PAWP: pulmonary artery wedge pressure, PVR: pulmonary vascular resistance, WU: Woods unit, PAC: pulmonary artery compliance, CO: cardiac output, VO<sub>2</sub>: oxygen consumption, V<sub>E</sub>/VCO<sub>2</sub>: ventilatory efficiency of carbon dioxide, OUES: oxygen uptake efficiency slope, EDVi: end-diastolic volume index, ESVi: end-systolic volume index, EF: ejection fraction, LA: left atrium.

In general, high-risk CpcPH patients were likely to be male and had increased LV mass and LA dilation on echocardiogram. Hemodynamics revealed lower cardiac index and pulmonary arterial compliance at rest and with exercise, and an increased mPAP/CO slope. Cardiopulmonary exercise data revealed decreased peak VO2 and OUES, and an increased VE/VCO2 slope. cMRI revealed an increased RV size, lower RVEF, decreased biventricular strain, and increased septal angle.

More interestingly, features associated with a low-risk CpcPH state were female sex, higher BMI, and lower LV mass and LA size on echocardiogram. On iCPET, mPAP/CO slope was lower, along with better aerobic capacity (VO2 and OUES) and ventilatory efficiency (VE/VCO2).

### 3.4 Pulmonary vascular mechanics (flow dynamics) among low- and high-risk phenogroups

Among the two defined phenogroups of PH-HFpEF (Table 1, Figure 1), unique patterns of pulmonary vascular wave mechanics metrics are shown in Figure 2 (and Table S2). The characteristic impedance (proximal PA stiffness) was significantly higher in high-risk PH-HFpEF phenogroup. Among metrics of WIA, forward compression wave in systole (ventricular energy expenditure and force generation) was also higher in high-risk phenogroup, which may indicate energy inefficiency.^45,57,60^ Among the decompression waves (aka. diastolic waves), RV relaxation is captured by forward wave, while distal pulmonary vascular reflections are captured by backward wave and diastolic reflection index (ratio of diastolic forward:backward wave).^58,63,64^ Diastolic reflection index revealed an interesting trend from rest-to-exercise. Although statistically insignificant, the high-risk phenogroup had worsening distal vascular reflections with exercise, while distal vascular reflections in low-risk PH-HFpEF and pre-capillary PH were either unchanged or decreased with exercise.

**Figure 2.**
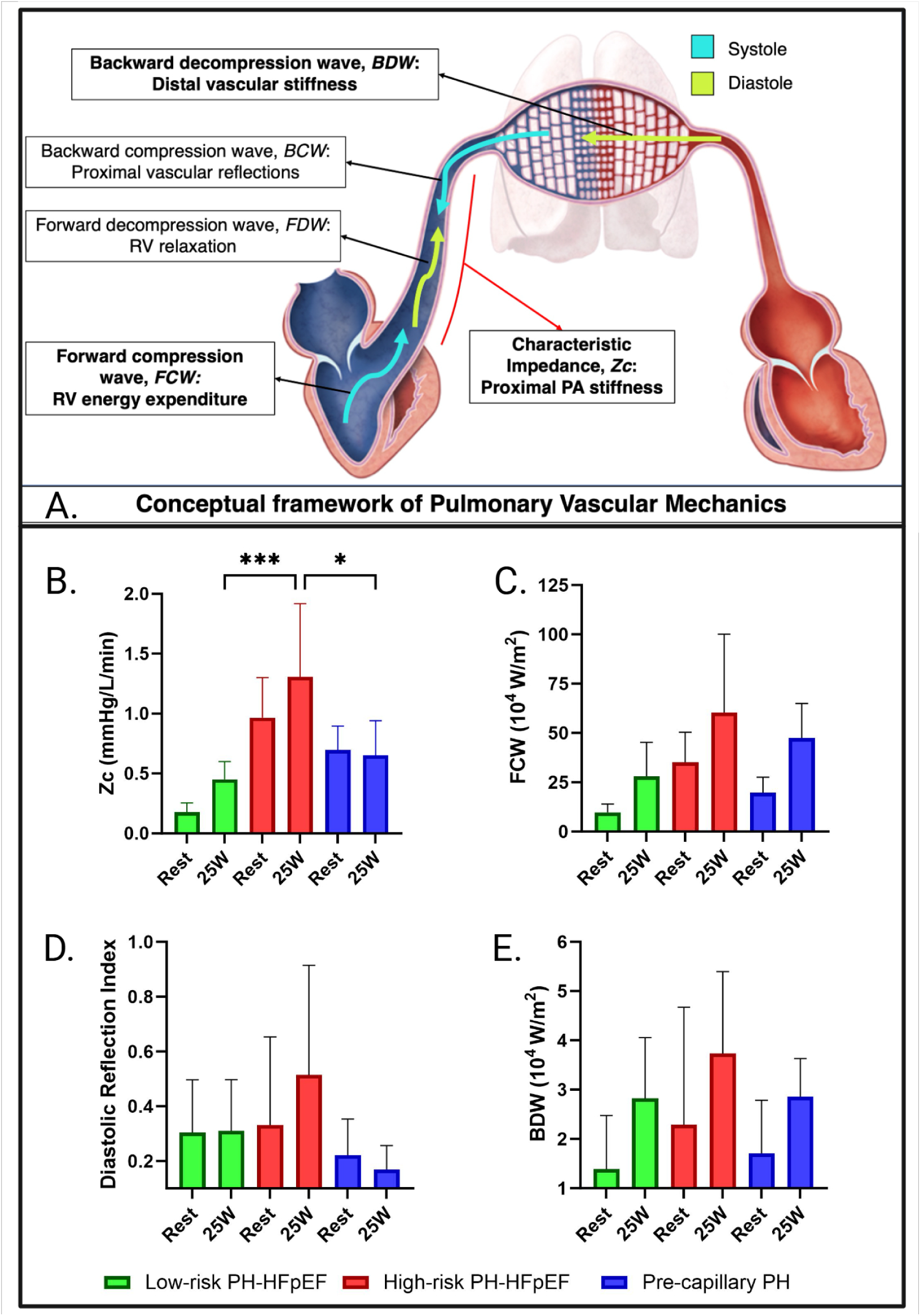
Invasive pulmonary vascular mechanics. Conceptual framework (A) and key metrics among PH-HFpEF phenogroups (low-risk vs high risk) and pre-capillary comparison group (B-E). Diastolic reflection index = backward decompression wave/forward decompression wave * p-value <0.05, *** p-value <0.001

### 3.5 4D flow MRI-based ventricular and pulmonary vascular mechanics (morphologic) among low- vs high-risk phenogroups

Among the two defined phenogroups of PH-HFpEF (Table 1, Figure 1), unique patterns of cardiac MRI and cardiopulmonary 4D flow metrics are shown in Figure 3 (and Table S3). The low-risk PH-HFpEF representative participant had a laminar flow, compared to disturbed flow in high-risk PH-HFpEF participant. RVEF was significantly higher in low-risk PH-HFpEF phenogroup, compared to high-risk phenogroup and pre-capillary PH. Although statistically non-significant, noticeable trends on 4D flow analyses revealed an increased viscous energy loss in main PA and left superior pulmonary vein in high-risk PH-HFpEF phenogroup^72–75^. Post-vasodilator 4D flow data in 6 PH-HFpEF participants (3 low- and 3 high-risk) showed feasibility of tracking 4D flow metrics post-vasodilator (Table S3). While statistically insignificant, there was a trend towards reduction in peak velocity in main PA and left superior pulmonary vein post-vasodilator in low-risk (possibly indicating vasoreactivity), compared to no change in peak velocities in high-risk participants.

**Figure 3.**
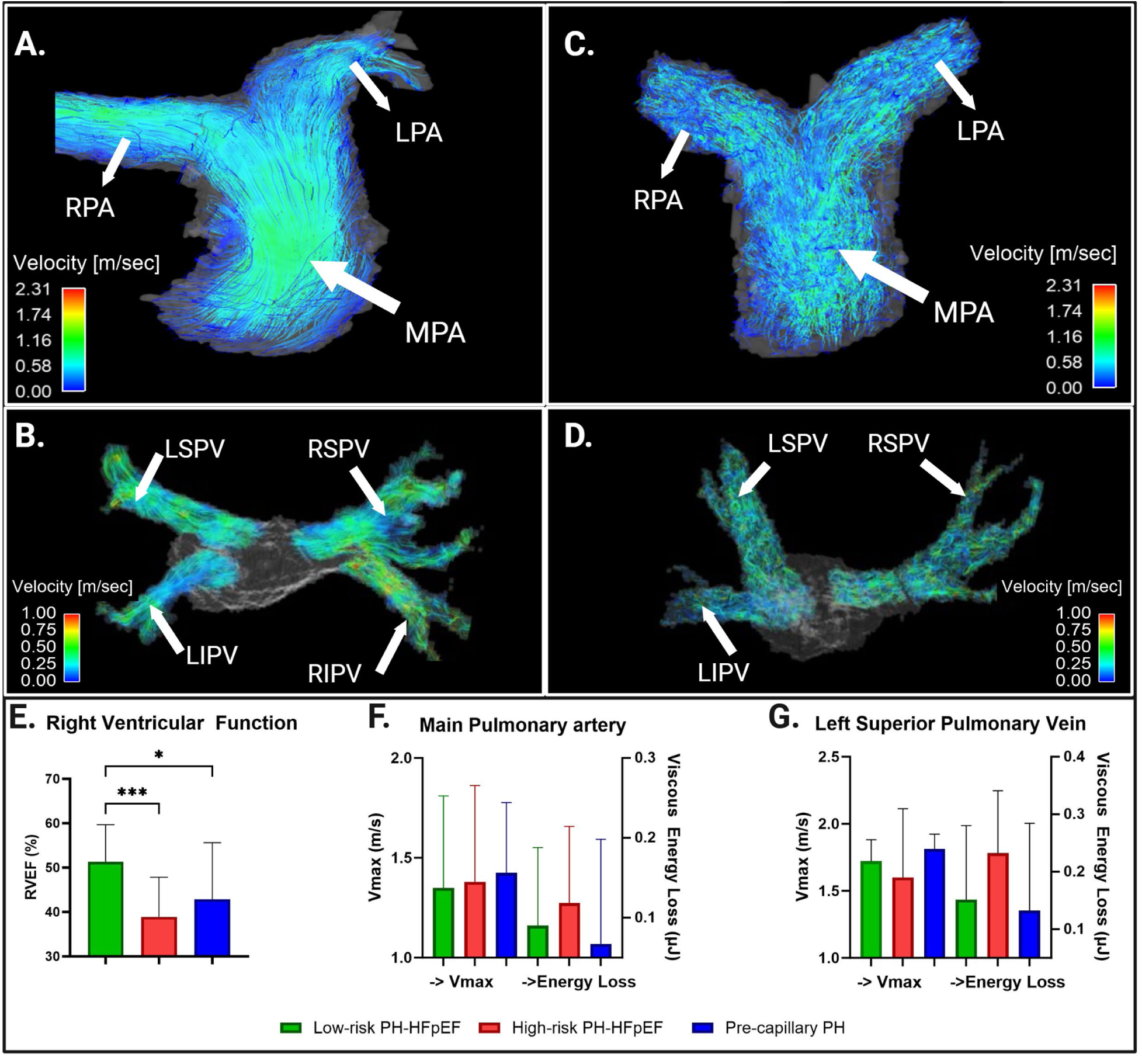
Integrative cardiac MRI with 4D flow. Representative qualitative comparison of 4D flow streamline visualizations in low-risk (A & B) vs. high-risk (C & D) PH-HFpEF, and key metrics among PH-HFpEF phenogroups (low-risk vs high risk) and pre-capillary comparison group (E-G). Streamline patterns in the low-risk group are characterized by long, continuous streamlines demonstrating laminar-like flow patterns as compared to the much shorter, disturbed and chaotic streamlines of the high-risk group demonstrating turbulent flow patterns.

### 3.6 Differential gene expression profiles distinguish high- and low-risk PH-HFpEF phenogroups

To uncover phenogroup-specific molecular signatures, endomyocardial biopsies from five patients each in the high- and low-risk PH-HFpEF groups (aggregate characteristics summarized in Table S4) were subjected to transcriptomic analysis using PacBio long-read RNA-seq technology (Figure 4A). The resulting data demonstrated high quality and consistency, with each sample yielding approximately 70 million total reads, over 98% of which were classified as polyadenylated. Mean read lengths were approximately 1,500 bp, and the number of unaligned reads was low, indicating robust alignment performance and overall data integrity (Table S5). PCA revealed clear segregation between the low- and high-risk phenogroups, reflecting distinct global transcriptomic profiles (Figure 4B).

**Figure 4.**
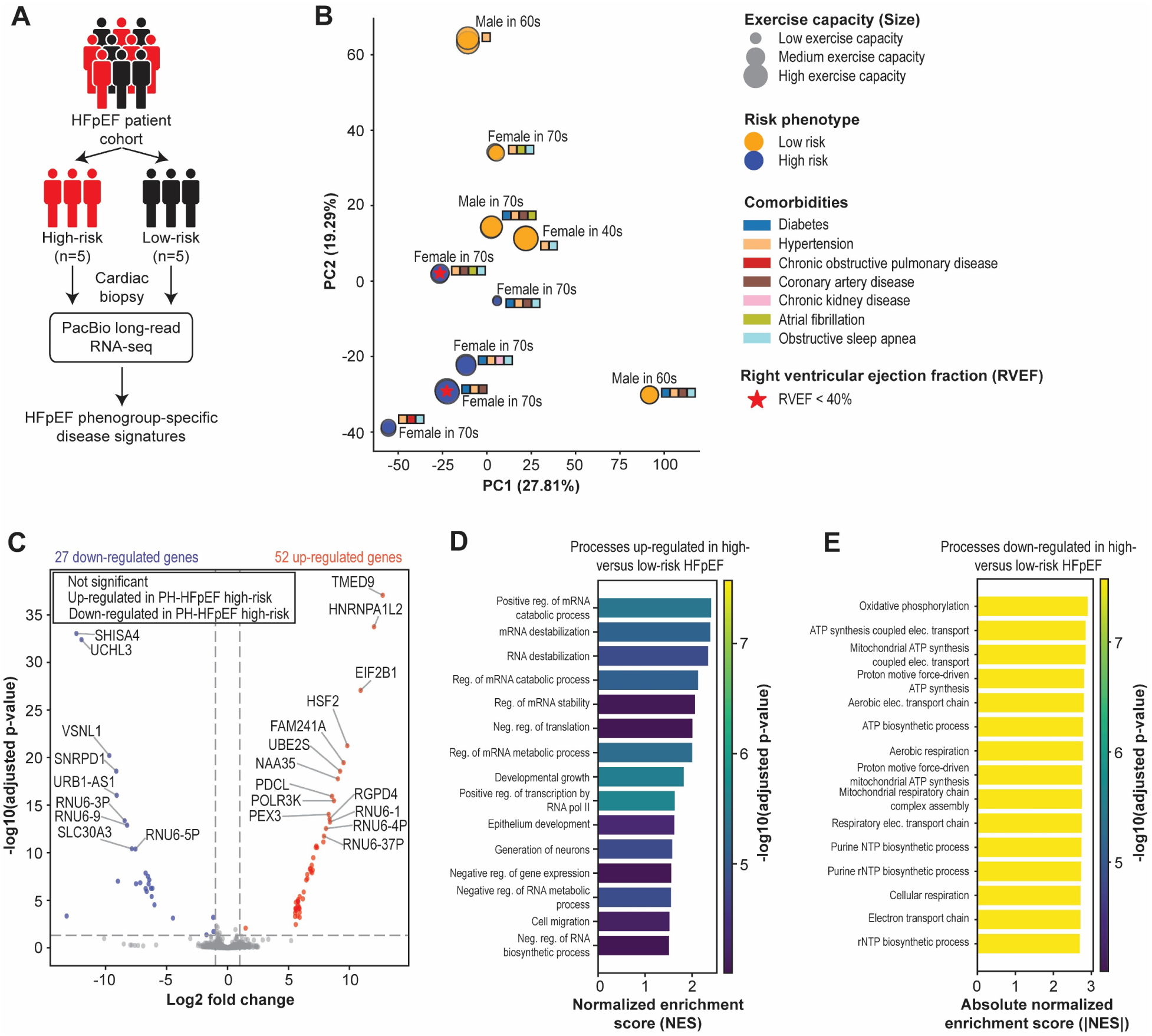
Long-read RNA-seq analysis of DEGs in the high-versus low-risk PH-HFpEF phenogroups. (A) Schematic overview of transcriptomic analysis of patients in the high- and low-risk phenogroups using third generation long-read RNA-seq. (B) Principal component analysis shows separation of individuals in the low- and high-risk PH-HFpEF groups. A technical replicate is present with each sample. (C) Volcano plot showing genes (among those with ≥10 TPM) up- and down-regulated in high- vs. low-risk PH-HFpEF groups. Bar plots showing biological process terms up- (D) and down-regulated (E) as determined by GSEA analysis in the high-risk PH-HFpEF phenogroup compared to the low-risk one.

Among the 12,561 genes with a mean count ≥10 in either group, 79 were differentially expressed (adjusted *p*< 0.05 and |log2fc| > 1). Of these, 52 genes were up-regulated and 27 were down-regulated in the high-risk group compared to the low-risk PH-HFpEF group (Figure 4C). GSEA of up-regulated genes in the high-risk PH-HFpEF phenogroup revealed enrichment of biological processes related to RNA metabolism and stability, as well as translational regulation, development, and gene expression pathways (Figure 4D). In contrast, GSEA of down-regulated genes in high-risk PH-HFpEF group identified enrichment of terms associated with mitochondrial function (Figure 4E), suggesting a potential link between mitochondrial dysfunction and increased clinical risk in PH-HFpEF.

### 3.7 Alternative transcript usage highlights post-transcriptional remodeling in high-risk PH-HFpEF

To further explore phenogroup-specific differences in post-transcriptional regulation, we leveraged the long-read RNA-seq data to assess alternative transcript usage in the myocardium of high- and low-risk PH-HFpEF patients. We compared log₂ fold changes in gene expression to the proportion of alternatively spliced transcripts contributing to total gene expression, revealing distinct transcriptomic signatures that differentiate the two phenogroups.

A subset of genes exhibited dominant alternative transcript usage, differentially skewed toward either the high-risk (red) or low-risk (blue) groups (Figure 5A), suggesting a potential regulatory layer modulating phenotypic divergence. Both phenogroups exhibited similar distributions of alternative splicing-event types. The most prevalent events included alternative polyadenylation, exon elongation greater than 50 bp, and intron retention (Figure 5B-C), indicating a shared repertoire of splicing mechanisms. Analysis of novel transcripts among the 4019 genes with transcripts ≥10 TPM revealed that the majority (78.6%, n=3160) expressed exclusively reference transcripts. Among the smaller subset with any novel isoform expression, the distribution of novel-transcript percentages was similar between high- and low-risk groups: counts tapered through the 1–25 %, 25-50% and 50–75 % bins and rose again in the 75–100 % bin, indicating a mix of modest and very high novel usage rather than a single dominant pattern (Figure 5D). GO enrichment analysis of alternatively spliced genes revealed strong association with biological processes relevant to muscle structure development, cytoskeleton organization, and circulatory system function (Figures 5E-F). These enrichments were consistent across both phenogroups and do not correlate with expression level, indicating conserved biological themes underlying alternative splicing activity (Figures 5E-F).

**Figure 5.**
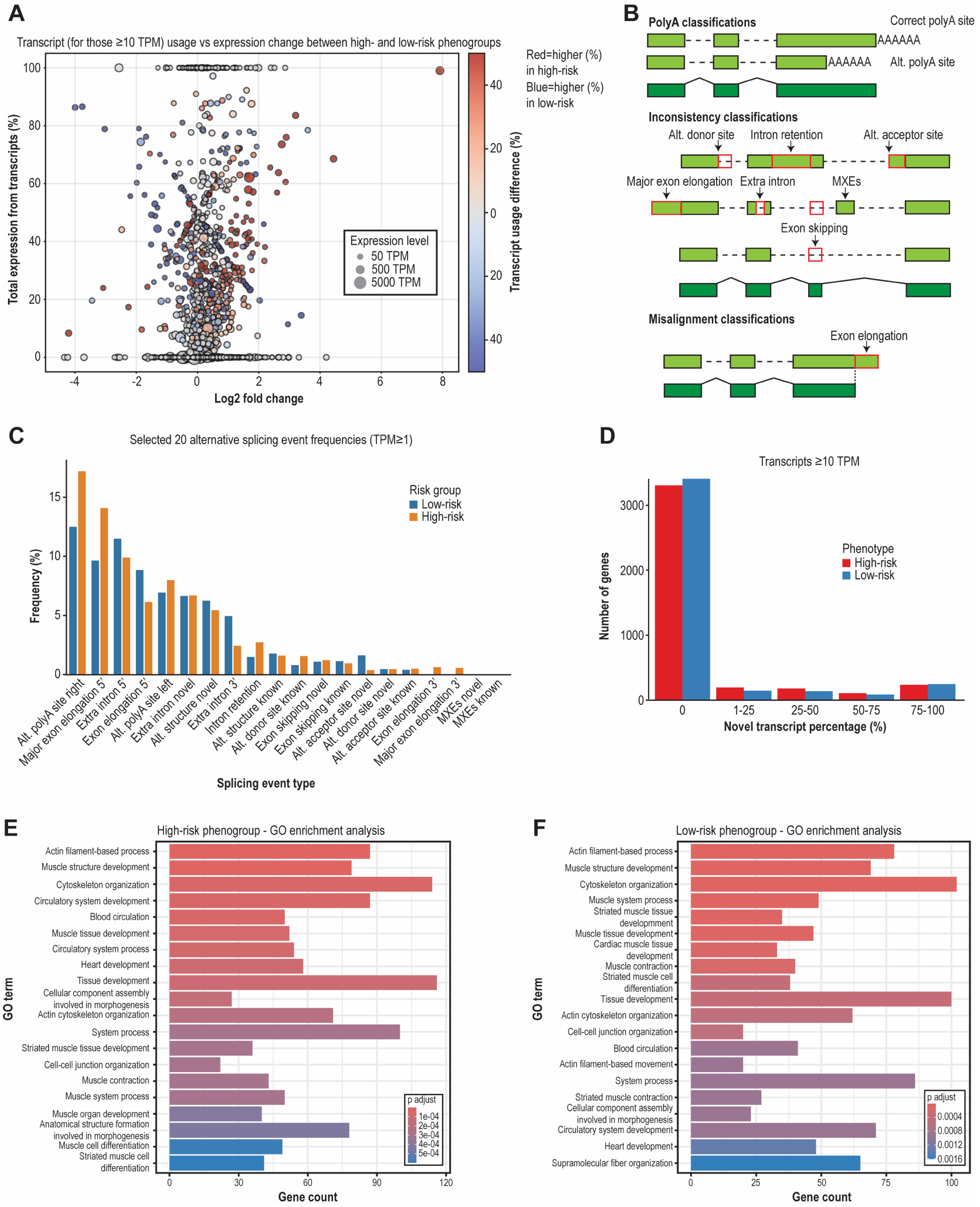
Long-read RNA-seq analysis of alternative transcript usage in high-versus low-risk PH-HFpEF phenogroups. (A) Scatter plot showing the log2 fold change of gene expression between high- and low-risk phenogroups (x-axis) and the percentage contribution of alternatively spliced transcripts to total gene expression (y-axis). The color scale indicates the predominant phenogroup of alternative splcing(red: high-risk dominance, blue: low-risk dominance), and dot size represents overall transcript expression levels (TPM). (B) Schematic illustrating different types of splicing events identified. (C) Bar graph showing the frequency of selected splicing events (TPM ≥ 1) observed in both high- and low-risk phenogroups. (D) Distribution of novel transcript percentages among the 4019 genes that contain transcripts with expression levels ≥ 10 TPM, comparing high- and low-risk phenogroups. (E-F) GO enrichment analysis of genes with alternatively spliced transcripts (≥10 TPM), highlighting biological processes enriched in high-risk (E) and low-risk (F) phenogroups, respectively.

In total, 5536 transcripts were differentially expressed (adjusted *p*< 0.05 and |log2fc| > 1). between the myocardium of individuals in the high-versus low-risk PH-HFpEF phenogroups, with 3212 upregulated and 2324 downregulated in the high-risk group (Figure 6A). Among these, several genes exhibiting distinct transcript usage were associated with biological processes previously implicated in HFpEF (Figure 6A)^76^. These include important regulators of hypertrophic signaling and signal transduction modulation (e.g., *ILK*), and mitochondrial metabolism and Ca^2+^-handling (*MT-CO1*, *MT-ND6*, and *VDAC3*) (Figure 6A).

**Figure 6.**
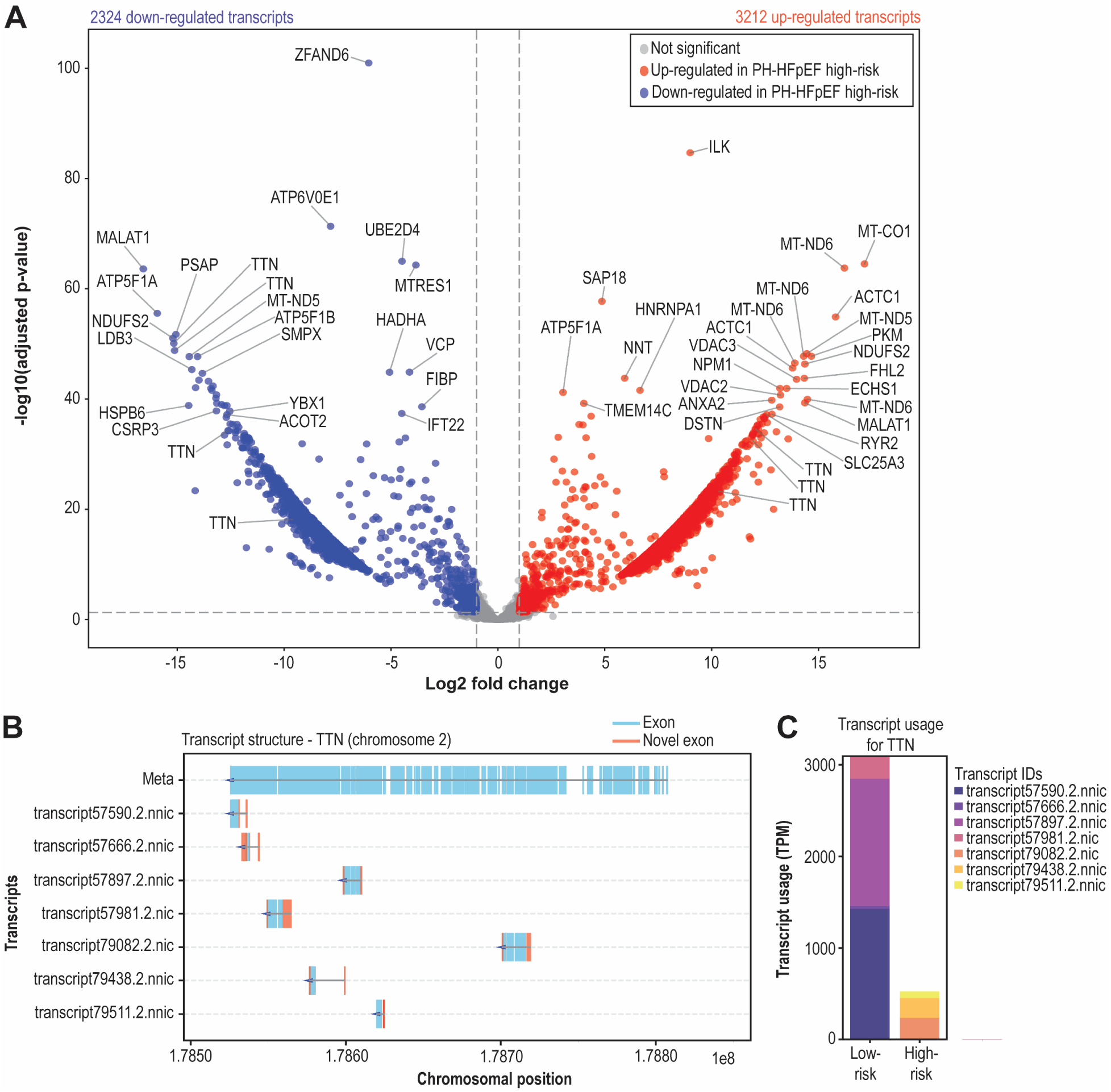
Transcript isoforms differentially expressed in high-versus low-risk PH-HFpEF phenogroups. (A) Volcano plot showing transcript isoforms up- and down-regulated in high-versus low-risk PH-HFpEF. Plot (B) and bar graph (C) showing *TTN* transcript and usage profiles in different phenogroups.

A notable finding was the differential usage of the *TTN* transcripts (Figure 6B-C and Table S6), which encode a giant sarcomeric protein titin that plays a determinant role in myocardial stiffness, a hallmark of HFpEF^77–80^. Long-read RNA-seq data enabled precise resolution of alternative transcript structures, revealing not only inclusion of canonical reference exons (shown in blue), but also novel exons (shown in red) that arise from mainly alternative structural configurations including alternative transcript start sites, non-canonical exon junctions, and alternative polyadenylation events (Figure 6B, Table S6). These structural variations result in novel transcript isoforms that do not match the annotated reference transcript splice sites (Figure 6B) and exhibit distinct expression patterns between high- and low-risk PH-HFpEF phenogroups (Figure 6C). These findings suggest that alternative transcript structures may contribute to the functional heterogeneity of titin isoforms, with implications for myocardial stiffness and disease progression in PH-HFpEF.

## 4. Discussion

In this study, we demonstrate that rich datasets can be leveraged to refine disease heterogeneity using unsupervised analytical methods to identify novel phenotypes in PH-HFpEF. Invasive pulmonary vascular mechanics and non-invasive 4D Flow cMRI methods build the basis to better understand and quantify in-depth myocardial and pulmonary vascular changes in these phenotypes. Lastly, we reported myocardial biopsy-based unique differences in these PH-HFpEF phenotypes, which shed light on biological mechanistic differences between low-risk vs high-risk disease phenogroups. These findings are a first step towards creating a geographic patient-specific map of abnormalities in pulmonary circulation-left heart axis and links to unique biological features, which can potentially lead to defining targetable abnormalities in vascular and ventricular mechanics.

Unsupervised machine learning methods have been shown to reveal significant insights into disease heterogeneity in HFpEF and PH.^81,82^ In this study, unsupervised K-means clustering approach provides powerful insights into specific sub-groups and phenotypes. To identify specific PH-HFpEF participants at risk of adverse clinical outcomes, the first clustering model using non-invasive clinical variables only, was comparable to standard-of-care invasive right heart catheterization-based PH-HFpEF groups (IpcPH vs CpcPH). The next two clustering analyses, adding exercise hemodynamics and then cMRI, enabled identification of a very high-risk PH-HFpEF phenogroup (Hazard ratio∼12). Morphologically, these high-risk participants had increased LV mass, poor LV strain and RV function. These findings are consistent with prior literature indicating poor RV function and RV:PA uncoupling as a high-risk feature in different cardiac-PVDs.^18,19,21,23,83,84^ Compared to low-risk PH-HFpEF phenogroup, these high-risk participants had similar PA pressures at rest, but worse exercise PH, CO augmentation, gas exchange and peak workload. More interestingly, a noticeable sex difference was found within the CpcPH group (Table 2), where there was an increased incidence of male sex in high-risk (vs low-risk) group. In other words, female sex with CpcPH was associated with a low-risk PH-HFpEF phenotype, compared to male sex.

These observed sex differences warrant further investigation to validate the findings and to elucidate the underlying biological mechanisms.

To characterize abnormal flow dynamics of PVD in PH-HFpEF, we employed models of invasive pulmonary vascular mechanics, which include characteristic impedance and wave intensity analyses. As a metric of proximal PA stiffness and direct contributor to RV afterload, characteristic impedance captures both load-dependent and structural-dependent mechanisms.^29,54–56^ Increased characteristic impedance in high-risk phenogroup indicates stiffer proximal pulmonary artery. Among wave intensity metrics, high-risk phenogroup had increased RV force generation (forward compression) and increased pulmonary vascular reflections (backward compression).^45,57,60^ Although statistically insignificant, high-risk phenogroup had increased distal vascular reflections in diastole (backward decompression wave and diastolic reflection index), which may indicate diseased distal vasculature^58,63,64^ (e.g., pulmonary venules, similar to prior reports of pulmonary venular hypertrophy in CpcPH^28^). These reflections are uniquely different from pre-capillary PH with a similar PVR. Overall, the flow dynamics model of pulmonary vascular mechanics demonstrates potential for identifying significant vessel-specific changes that are unique to distinct PH subtypes.

To characterize abnormal morphologic changes of PVD in PH-HFpEF, we reported the cMRI-based strain, mass, and volume data of cardiac chambers. However, we reported 4D flow MRI-based flow metrics and their vasoreactivity, which include peak velocity, viscous energy loss and kinetic energy. Although statistically insignificant, PA and veins in high-risk PH-HFpEF phenogroup had trend towards increased viscous energy loss and kinetic energy (indicating energy inefficiency), along with poor vasoreactivity to nitroglycerine (indicating structural changes and fixed stiffness of PA and veins). These findings are preliminary, yet they provide a framework for future studies to localize changes across pulmonary circulation-left heart axis using novel metrics like PA distensibility, sheer wall stress, viscous energy loss, and kinetic energy efficiency.^46,53,72^

Although prior studies have identified distinct HFpEF phenogroups based on clinical characteristics and differential responses to therapies,^81,85–87^ few have investigated the molecular features that distinguish these subgroups, and none have assessed differences at the post-transcriptional level. In this study, we employed third generation long-read RNA-seq for the comprehensive transcriptomic profiling of myocardial biopsies from patients with high-versus low-risk PH-HFpEF. This study aimed to uncover molecular features associated with disease severity. Our data revealed that genes that were up- or down-regulated between the two phenogroups were enriched in pathways predominantly related to mRNA turnover/stability (upregulated in high-risk) and mitochondrial metabolic processes (downregulated in high-risk). These findings align with growing evidence that post-transcriptional RNA regulation and mitochondrial dysfunction are central features of heart failure pathogenesis, particularly in HFpEF. Consistent with prior reports implicating aberrant pre-mRNA splicing in heart failure,^88–90^ we observed multiple genes encoding core spliceosome components (e.g., *HNRNPA1L2*, *SNRPD1*, *RNU6-3P*, *RNU6-1*, *RNU6-9*, etc.) that were differentially expressed. Additionally, several translational initiation factors (i.e., *EIF2B1* and *EIF1AY*) were also among the most differentially expressed genes, suggesting that altered post-transcriptional RNA processing and translational control may contribute to phenotypic divergence between these HFpEF subgroups. These transcriptional signatures likely reflect a cellular stress response aimed at adapting to increased demand or injury but may ultimately drive maladaptive remodeling in the myocardium.

Importantly, the use of long-read RNA-seq allowed us to assess differential transcript usage, providing insights into isoform-level changes not easily captured by short-read technologies. Notably, approximately 80% of transcripts (≥10 TPM) corresponded to reference isoforms, indicating that most transcript-level differences were driven by quantitative changes in expression rather than widespread generation of novel isoforms. However, alterations in alternative polyadenylation, exon elongation, and intron retention were still observed and could contribute to functional diversity and regulation in these phenogroups. Despite the relatively modest prevalence of novel isoforms, genes with non-zero alternative splicing rates disproportionately affect pathways adjacent to HFpEF pathogenesis. Most notably, we observed phenogroup-specific splicing of *TTN*, which encodes the titin protein.^91,92^ Given titin’s well-established role in regulating myocardial passive stiffness—a hallmark of HFpEF—and its known involvement in diastolic dysfunction,^77–80,93–95^ our findings suggest that titin isoform imbalance may be a key molecular feature distinguishing low-from high-risk PH-HFpEF. This highlights *TTN* splicing as a potential therapeutic target in addressing myocardial stiffness and disease progression in this patient population.

The current study advances the field in three specific ways. First, deep morphologic and exercise hemodynamics-based characterization defined a high-risk PH-HFpEF phenogroup with worse PH and RV function. Second, and more importantly, we have reported a framework (flow dynamics and morphologic cMRI) of in-depth characterization of pulmonary vascular disease and a pipeline to link it with biological myocardial changes. Third, for the first time, transcriptomic profiling implicated differences in post-transcriptional RNA processing in disparate clinical outcomes in PH-HFpEF. In the future, larger studies are needed to define biological mechanisms that link to specific segments of pulmonary vascular bed, which are studied at rest and under stress (physiological stress of exercise and/or vasodilator) to define structural/fixed vs vasoreactive changes.

## 5. Limitations

This is a single center study with a modest sample size, albeit this is not uncommon with a highly specialized study protocol. The methods and findings reported here may require validation in larger, multi-center cohorts. Moreover, while we observed intriguing sex differences, these findings also need to be carefully validated in larger human studies with balanced sex distribution and further mechanistic studies need to be defined in well-characterized animal models. Despite these limitations, our study provides novel and important insights into subtypes of HFpEF and highlights the potential molecular targets for precise medicine. Future studies will involve collaboration with other centers to validate and expand upon these findings.

## 6. Conclusions

In a cohort of 42 PH-HFpEF participants with deeply phenotyped data including invasive exercise hemodynamics and cMRI, unsupervised clustering methods revealed two distinct phenogroups (clusters). The clinical outcomes varied significantly among these two phenogroup (labeled as high-risk and low-risk, with a Hazard ratio=11.96 for adverse outcomes in one-year). High-risk phenogroup had increased LV mass, poor LV strain, poor RV strain and decreased RVEF. Additionally, high-risk phenogroup had worse exercise PH and gas exchange. Vascular mechanics models of flow dynamics and 4D flow MRI revealed increased proximal PA stiffness (characteristic impedance), RV force generation (energy inefficiency), increased distal vascular reflections with exercise (diastolic reflection index), and increased energy loss in PA and vein, in high-risk PH-HFpEF phenogroup. Lastly, gene and transcript usage profiles suggest that changes in post-transcriptional RNA processing accompany evolution from the low- (normal RV) to high-risk (RV failure) disease states.

## Data Availability

All data produced in the present study are available upon reasonable request to the authors

## Non-standard Abbreviations and Acronyms

HFpEF: heart failure with preserved ejection fraction
PH: pulmonary hypertension
PVD: pulmonary vascular disease
RV: right ventricular
PVR: pulmonary vascular resistance
IpcPH: isolated post-capillary PH
CpcPH: combined pre-/post-capillary PH
cMRI: cardiac MRI
RNA-seq: RNA-sequencing
iCPET: invasive cardiopulmonary exercise testing
LV: left ventricular
EF: ejection fraction
PA: pulmonary artery
VO2: peak oxygen consumption
VE/VCO2: slope of minute ventilation/carbon dioxide production
ETCO2: end-tidal carbon dioxide from rest-to-exercise
LA: left atrium
WIA: wave intensity analysis
PCA: principal component analysis
GSEA: gene set enrichment analysis
GO: gene ontology
ANOVA: analysis of variance
mPAP/CO: mean pulmonary arterial pressure/cardiac output
OUES: oxygen uptake efficiency slope

## Acknowledgments

None

