## Supplemental Data for "Multimodal Characterization of High-risk PH-HFpEF phenogroup with Right Ventricular Dysfunction: Vascular Mechanics and Myocardial Transcriptomics"

**SUPPLEMENTAL DATA**
**Methods: Wave Intensity Analysis**

**Figure S1. Myocardial fibrosis quantification in a PH-HFpEF patient: mild fibrosis on endomyocardial biopsy and increased fibrosis metrics on cardiac MRI (increased T1 time and extracellular volume)**

**Table S1. Variables used in clustering analyses**
**Table S2. Features of K-means clusters: Wave Mechanics**
**Table S3. Features of K-means clusters: 4D flow MRI**
**Table S4. Features of K-means clusters: Myocardial biopsy data**
**Table S5. Sequencing and alignment metrics from PH-HFpEF risk groups**
**Table S6. Titin transcript features**

### Methods: Wave Intensity Analysis

#### Analysis of Wave Speed, Wave Intensity, and Wave Separation

From continuously obtained, time-dependent, PAP and U waveforms, 4 to 15 PAP waveforms and 2 to 3 U waveforms were selected that were free from motion artifacts and ensemble-averaged to create representative single-beat waveforms. The early systolic segment of pressure was then aligned with the early systolic segment of velocity, creating PU loops for wave intensity analyses. Wave speed $c$ (in m/s) was calculated using:

$$c = \frac{1}{\rho}\sqrt{\frac{\sum dP^2}{\sum dU^2}}$$

where ρ is the density of blood (assumed equal to 1040 kg/m$^3$), dP is the point-to-point change in the single-beat PAP waveform over an interval dt (in mmHg), and dU is the change in the single-beat velocity waveform over the same interval dt (in m/s)[57,96]. Then waves were separated into their forward and backward components via the following equation:

$$WI_{\pm} = \left(\frac{\text{dP x CCD}}{\text{dt}} \pm \frac{\text{dU x CCD}}{\text{dt}}\right)^2/(4\rho c)$$

where dP is the change in pressure, dU is the change in velocity, dt is the time interval, CCD is the cardiac cycle duration (inverse of heart rate) [57,96]. $WI_+$ indicates wave propagation in the forward direction in the PA (away from the RV), and $WI_-$ indicates wave propagation in the backward direction in the PA (towards the RV) [57,96]. The waves were further defined as compression (i.e., increasing pressure) or decompression (i.e., decreasing pressure) components. FCWs were characterized by an increase in both pressure and velocity, while BCWs were characterized by an increase in pressure but a decrease in velocity. FDWs were characterized by a decrease in both pressure and velocity, while BDWs were characterized by a decrease in pressure but an increase in velocity[57,96].

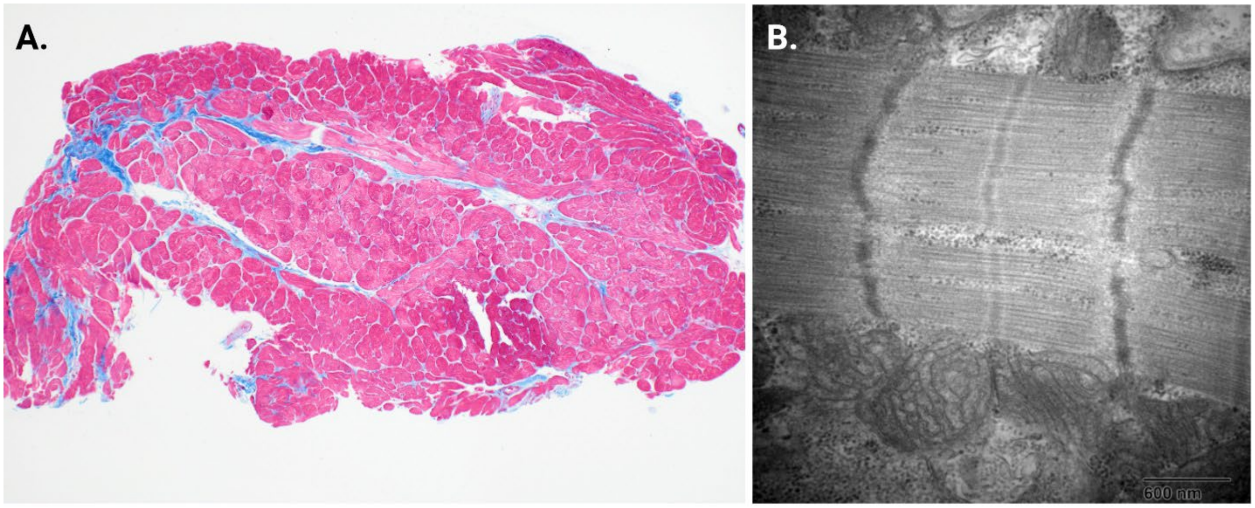


Male in their 60s with obesity (BMI=32), hypertension and diabetes

Endomyocardial tissue with mild myocyte hypertrophy and minimal interstitial fibrosis. Trichrome stain (**A**) showing fibrosis (blue) in approximately 15-20% of the tissue (fibrosis score=2). Electron microscopy **(B)** showing intact sarcomeres with normal distribution of cytoplasmic glycogen and normal mitochondria.

Cardiac MRI revealed left ventricular ejection fraction=65.3% and right ventricular ejection fraction=44.7%

Native myocardial T1 time: mildly elevated at 1218 m.sec and extra-cellular volume=31% (Normal: 25-28) with hematocrit=41

**Figure S1. Myocardial fibrosis quantification in a PH-HFpEF patient: mild fibrosis on endomyocardial biopsy and increased fibrosis metrics on cardiac MRI (increased T1 time and extracellular volume)**

**Table S1. Variables used in clustering analyses.**

| **Clinical Variables** | **Echocardiographic Variables** | **Rest Hemodynamic Variables** | **Exercise Hemodynamic Variables** | **Cardiac MRI variables** |
|---|---|---|---|---|
| Age | Interventricular septal diameter in diastole | heart rate | heart rate | left ventricular mass |
| Sex 1-male, 2-female | Left ventricular internal diameter in diastole | systolic blood pressure | systolic blood pressure | left ventricular end-systolic volume indexed to body surface area |
| Race 1-African American A 2-White 3-Hispanic 4-other | Left ventricular posterior wall thickness in diastole | diastolic blood pressure | diastolic blood pressure | left ventricular end-diastolic volume indexed to body surface area |
| Smoking status | Eccentricity index in systole | mean arterial blood pressure | mean arterial blood pressure | right ventricular end-systolic volume indexed to body surface area |
| Alcohol use | right:left venticular ratio | right atrial pressure | right atrial pressure | right ventricular end-diastolic volume indexed to body surface area |
| Hypertension | left atrial volume biplane index | Right ventricular systolic pressure | Systolic pulmonary arterial pressure | left ventricular mass index |
| Chronic obstructive pulmonary disease | right ventricular base/apex ratio in systole | Right ventricular end-diastolic pressure | Diastolic pulmonary arterial pressure | left ventricular strain longitudinal |
| Diabetes | Right ventricular fractional area change | Systolic pulmonary arterial pressure | Mean pulmonary arterial pressure | left ventricular strain cirumferential |
| Coronary artery disease | tricuspid anular plane systolic excursion | Diastolic pulmonar | Mean pulmonary arterial wedge pressure | left ventricular strain radial |

| | | | | |
|---|---|---|---|---|
| | | y arterial pressure | | |
| Chronic kidney disease | Mitral inflow Early diastolic velocity € | Mean pulmonar y arterial pressure | Transpulmonary gradient | right ventricular strain longitudinal |
| End-stage renal disease | Early diatolic mitral annulus velocity (e') medial | Mean pulmonar y arterial wedge pressure | Aortic saturation | right ventricular strain cirumferential |
| Interstitial lung disease | E/e' medial | Transpul monary gradient | Pulmonary artery saturation | right ventricular strain radial |
| Liver Cirrhosis | Early diatolic mitral annulus velocity lateral | Aortic saturatio n | Fick cardiac output | left ventricular strain rate Circumferential |
| Atrial fibrillation | E/e' lateral | Pulmonar y artery saturatio n | Fick cardiac index | left ventricular strain rate Longitudinal |
| Pulmonary Embolism | E/e' Average | Fick cardiac output | Pulmonary vascular resistance | left ventricular strain rate Radial |
| Connective Tissue Disease | pulmonary artery systolic pressure | Fick cardiac index | Systemic vascular resistance | right ventricular strain rate Circumferential |
| Obstructive sleep apnea | tricuspid anular plane systolic excursion/pulmonary artery systolic pressure | Pulmonar y vascular resistanc e | Stroke volume | right ventricular strain rate Longitudinal |
| Hemoglobin | Left ventricular ejection fraction | Systemic vascular resistanc e | Stroke volume index | right ventricular strain rate Radial |
| Creatinine | tricuspid anular plane systolic excursion normalized by right ventricular area in diastole | Stroke volume | Pulmonary arterial compliance | left ventricular time to peak strain Circumferential |
| NewYork Heart Association Status | tricuspid anular plane systolic excursion normalized by right | Stroke volume index | Percent change in pulmonary vascular | left ventricular time to peak strain Longitudinal |

| | | | | |
|---|---|---|---|---|
| | ventricular area in systole | | resistance from rest to exericse | |
| Pulmonary Hypertension vasodilator medication | left ventricular mass index | Pulmonary arterial compliance | Pulmonary arterial compliance | left ventricular time to peak strain Radial |
| Anticoagulant | | | Mean pulmonary arterial pressure/cardiac output slope | right ventricular time to peak strain Circumferential |
| Diuretic | | | pulmonary artery wedge pressure/cardiac output slope | right ventricular time to peak strain Longitudinal |
| Mineralocorticoid receptor antagonist | | | Peak watts | right ventricular time to peak strain Radial |
| Height (cm) | | | Rest end-tidal carbon dioxide | left atrial longitudinal strain |
| Weight (kg) | | | Exercise end-tidal carbon dioxide | left atrial strain rate |
| Body mass index | | | Minute Ventilation to carbon dioxide production slope | right atrial longitudinal strain |
| | | | Oxygen uptake efficiency slope | right atrial strain rate |
| | | | Peak heart rate | left atrial ejection fraction |
| | | | Peak oxygen pulse | right atrial ejection fraction |
| | | | peak respiratory exchange ratio | left atrial end-systolic volume indexed to body surface area |
| | | | peak oxygen consumption (ml/kg/min) | left atrial end-diastolic volume indexed to body surface area |
| | | | peak oxygen consumption, %predicted | right atrial end-systolic volume indexed to body surface area |

| | | | | |
|---|---|---|---|---|
| | | | | right atrial end-diastolic volume indexed to body surface area |
| | | | | epicardial fat |
| | | | | septal angle |
| | | | | right ventricular ejection fraction |
| | | | | left ventricular ejection fraction |

**Table S2. Vascular Mechanics with provocative maneuver (exercise) for vessel specific changes.**

| | | Low risk PH-HFpEF, n=11 | High-risk PH-HFpEF, n=6 | Pre-capillary PH, n=5 | p-value | | |
|---|---|---|---|---|---|---|---|
| | | | | | Low vs high | Low vs Pre | High vs Pre |
| **Ipc/CpcPH** | | **10/1** | **1/5** | **-** | | | |
| **Rest Hemodynamics:** | | | | | | | |
| mPAP (mmHg) | | 26.1±6.6 | 39.3±7.5 | 31.2±7.5 | 0.0009 | 0.094 | 0.05 |
| PAWP (mmHg) | | 16.2±3.9 | 19.3±3.0 | 12.8±1.6 | 0.053 | 0.043 | 0.001 |
| Cardiac output (L/min) | | 6.3±1.5 | 4.5±1.4 | 4.7±1.2 | 0.012 | 0.026 | 0.405 |
| PVR (Woods unit) | | 1.6±0.4 | 4.9±2.4 | 4.1±2.1 | 0.0002 | 0.0007 | 0.288 |
| PAC (mL/mmHg) | | 5.5±1.6 | 2.0±1.0 | 3.8±1.3 | 0.0001 | 0.011 | 0.049 |
| **Cardiopulmonary Exercise Test** | | | | | | | |
| Peak $VO_2$ (mL/Kg/min) | | 18.2±4.6 | 9.5±2.9 | 12.4±5.1 | 0.0004 | 0.019 | 0.135 |
| $V_E/VCO_2$ slope | | 31.7±6.6 | 40.6±10.7 | 38.9±9.1 | 0.027 | 0.049 | 0.392 |
| OUES (mL/min/Log[L/min]) | | 1.63±0.51 | 1.24±0.49 | 1.11±0.32 | 0.095 | 0.031 | 0.308 |
| **Zc** (mmHg/L.min$^{-1}$) | **Rest** | 0.18±0.08 | 0.97±0.33 | 0.69±0.19 | <0.0001 | <0.0001 | 0.074 |
| | **25watts** | 0.45±0.15 | 1.31±0.61 | 0.65±0.29 | 0.0002 | 0.041 | 0.028 |
| **Wave speed** (m/s) | **Rest** | 2.4±0.7 | 4.8±2.2 | 3.8±1.8 | 0.002 | 0.017 | 0.219 |
| | **25watts** | 3.2±1.2 | 4.7±2.1 | 4.4±1.5 | 0.037 | 0.046 | 0.406 |
| **Ees/Ea** | **Rest** | 1.12±0.34 | 0.62±0.25 | 0.58±0.22 | 0.003 | 0.003 | 0.406 |
| | **25watts** | 0.59±0.23 | 0.56±0.26 | 0.61±0.12 | 0.365 | 0.456 | 0.336 |
| **Ees** (mmHg/mL) | **Rest** | 0.27±0.12 | 0.58±0.52 | 0.27±0.09 | 0.035 | 0.494 | 0.112 |
| | **25watts** | 0.24±0.11 | 0.62±0.53 | 0.37±0.08 | 0.015 | 0.013 | 0.162 |
| **FCW** ($10^4$.W/m$^2$.s$^2$) | **Rest** | 9.8±4.2 | 35.1±15.2 | 17.8±7.8 | <0.0001 | 0.002 | 0.036 |
| | **25watts** | 28.1±17.1 | 60.4±39.7 | 47.4±17.6 | 0.015 | 0.028 | 0.259 |

| | | | | | | | |
|---|---|---|---|---|---|---|---|
| **BCW** ($10^4$.W/$m^2$.$s^2$) | **Rest** | 3.1±1.9 | 11.7±6.4 | 6.1±6.8 | 0.0003 | 0.092 | 0.095 |
| | **25watts** | 4.2±2.7 | 15.1±3.6 | 9.4±3.1 | <0.0001 | 0.0019 | 0.011 |
| **FDW** ($10^4$.W/$m^2$.$s^2$) | **Rest** | 4.7±2.5 | 6.6±2.1 | 9.3±7.2 | 0.069 | 0.038 | 0.206 |
| | **25watts** | 9.9±5.6 | 12.0±9.0 | 20.5±12.4 | 0.283 | 0.015 | 0.111 |
| **BDW** ($10^4$.W/$m^2$.$s^2$) | **Rest** | 1.4±1.1 | 2.3±2.4 | 1.7±1.1 | 0.148 | 0.297 | 0.314 |
| | **25watts** | 2.8±1.2 | 3.7±1.7 | 2.8±0.8 | 0.109 | 0.479 | 0.155 |
| **Systolic Reflection Index** | **Rest** | 0.32±0.13 | 0.34±0.17 | 0.26±0.19 | 0.388 | 0.227 | 0.233 |
| | **25watts** | 0.15±0.04 | 0.32±0.17 | 0.21±0.07 | 0.003 | 0.026 | 0.097 |
| **Diastolic Reflection Index** | **Rest** | 0.30±0.19 | 0.33±0.32 | 0.22±0.13 | 0.412 | 0.203 | 0.248 |
| | **25watts** | 0.31±0.19 | 0.51±0.40 | 0.17±0.09 | 0.083 | 0.069 | 0.047 |
| **Distensibility,** α | | 0.76±0.18 | 0.65±0.37 | 0.49±0.17 | 0.201 | 0.008 | 0.213 |

ABBREVIATIONS: PH-HFpEF: pulmonary hypertension due to heart failure with preserved ejection fraction, IpcPH: isolated post-capillary PH, CpcPH: combined pre-/post-capillary PH, mPAP: mean pulmonary artery pressure, PAWP: pulmonary artery wedge pressure, PVR: pulmonary vascular resistance, PAC: pulmonary artery compliance, $VO_2$: oxygen consumption, $V_E/VCO_2$: ventilatory efficiency of carbon dioxide, OUES: oxygen uptake efficiency slope, Zc: characteristic Impedance, FCW: forward compression waves, FDW: forward decompression waves, BCW: backward compression waves; BDW: backward decompression waves.

**Table S3. 4D flow MRI cardiopulmonary MRI data with provocative maneuver (post-vasodilator- sublingual nitroglycerine) for vessel specific changes.**

| | | **Low risk PH-HFpEF** | | **High-risk PH-HFpEF** | | **Pre-capillary PH, n=5** | **p-value** | | | |
|---|---|---|---|---|---|---|---|---|---|---|
| | | **Rest, n=6** | **ΔPost-Vasodilator, n=3** | **Rest, n=4** | **ΔPost-Vasodilator, n=3** | | **Low vs high** | **Low vs Pre** | **High vs Pre** | **Post-VD** |
| **Ipc/CpcPH** | | 4/2 | | 2/2 | | - | - | - | - | - |
| **Rest Hemodynamics:** | | | | | | | | | | |
| mPAP (mmHg) | | 29.7±11.7 | | 32.2±11.1 | | 31.5±8.8 | 0.37 | 0.40 | 0.46 | |
| PAWP (mmHg) | | 19.0±2.8 | | 17.0±2.0 | | 12.2±1.3 | 0.13 | <0.01 | <0.01 | |
| Cardiac output (L/min) | | 6.1±1.2 | | 5.4±1.7 | | 4.2±0.6 | 0.24 | 0.12 | 0.02 | |
| PVR (Woods unit) | | 2.1±0.7 | | 3.6±3.7 | | 4.6±2.2 | 0.18 | 0.02 | 0.33 | |
| PAC (mL/mmHg) | | 5.2±1.6 | | 3.0±1.9 | | 2.4±0.9 | 0.05 | 0.01 | 0.30 | |
| **Cardiopulmonary Exercise Test:** | | | | | | | | | | |
| Peak $VO_2$ (mL/Kg/min) | | 18.9±1.3 | | 16.1±5.1 | | 14.9±7.1 | 0.16 | 0.39 | 0.16 | |
| $V_E/VCO_2$ slope | | 30.1±7.3 | | 40.4±7.5 | | 39.6±10.4 | 0.05 | 0.45 | 0.12 | |
| OUES (mL/min/Log[L/min]) | | 1.85±0.98 | | 1.28±0.49 | | 1.04±0.33 | 0.17 | 0.22 | 0.09 | |
| **Pulmonary Artery** | **Energy Loss** (µJ) | 0.09±0.09 | 0.10±0.06 | 0.12±0.09 | 0.15±0.04 | 0.07±0.13 | 0.33 | 0.37 | 0.27 | 0.33 |
| | **Kinetic Energy** (µJ) | 0.09±0.05 | 0.10±0.07 | 0.12±0.08 | 0.15±0.06 | 0.08±0.05 | 0.22 | 0.35 | 0.16 | 0.20 |
| | **Peak Velocity** (m/sec) | 1.3±0.5 | 1.2±0.6 | 1.4±0.5 | 1.5±0.5 | 1.4±0.3 | 0.46 | 0.38 | 0.43 | 0.28 |
| **Pulmonary** | **Energy** | 0.14±0.14 | 0.15±0.16 | 0.19±0.16 | 0.21±0.11 | 0.08±0.11 | 0.31 | 0.28 | 0.16 | 0.29 |

| | | | | | | | | | |
|---|---|---|---|---|---|---|---|---|---|
| **vein (right superior)** | **Loss** (µJ) | | | | | | | | |
| | **Kinetic Energy** (µJ) | 0.11±0.06 | 0.12±0.07 | 0.15±0.11 | 0.18±0.10 | 0.10±0.04 | 0.25 | 0.38 | 0.21 | 0.25 |
| | **Peak Velocity** (m/sec) | 1.7±0.1 | 1.8±0.1 | 1.6±0.4 | 1.6±0.5 | 1.7±0.1 | 0.27 | 0.44 | 0.31 | 0.36 |
| **Pulmonary vein (left superior)** | **Energy Loss** (µJ) | 0.15±0.13 | 0.09±0.11 | 0.23±0.11 | 0.22±0.10 | 0.13±0.15 | 0.19 | 0.42 | 0.20 | 0.13 |
| | **Kinetic Energy** (µJ) | 0.11±0.05 | 0.11±0.12 | 0.18±0.09 | 0.18±0.09 | 0.12±0.07 | 0.09 | 0.39 | 0.22 | 0.28 |
| | **Peak Velocity** (m/sec) | 1.7±0.2 | 1.5±0.4 | 1.6±0.5 | 1.6±0.5 | 1.8±0.1 | 0.29 | 0.20 | 0.26 | 0.45 |
| **Aorta** | **Energy Loss** (µJ) | 0.14±0.14 | 0.15±0.14 | 0.17±0.14 | 0.20±0.08 | 0.11±0.12 | 0.36 | 0.40 | 0.28 | 0.33 |
| | **Kinetic Energy** (µJ) | 0.11±0.06 | 0.13±0.09 | 0.14±0.09 | 0.16±0.8 | 0.12±0.03 | 0.26 | 0.13 | 0.39 | 0.38 |
| | **Peak Velocity** (m/sec) | 1.5±0.4 | 1.5±0.5 | 1.6±0.4 | 1.6±0.5 | 1.7±0.1 | 0.47 | 0.15 | 0.19 | 0.42 |

ABBREVIATIONS: PH-HFpEF: pulmonary hypertension due to heart failure with preserved ejection fraction, IpcPH: isolated post-capillary PH, CpcPH: combined pre-/post-capillary PH, mPAP: mean pulmonary artery pressure, PAWP: pulmonary artery wedge pressure, PVR: pulmonary vascular resistance, PAC: pulmonary artery compliance, $VO_2$: oxygen consumption, $V_E/VCO_2$: ventilatory efficiency of carbon dioxide, OUES: oxygen uptake efficiency slope.

**Table S4. Characteristics of PH-HFpEF participants undergoing myocardial biopsy and RNA-seq.**

| | **Low risk**<br>**N=5** | **High risk**<br>**N=5** |
|---|---|---|
| **IpcPH/CpcPH** | 3/2 | 0/5 |
| **Age** (years) | 67.2±11.8 | 74.4±2.9 |
| **Female, n** | 2 | 5 |
| **BMI** (Kg/m$^2$) | 35.3±3.1 | 37.0±6.3 |
| **Comorbidities, n** | | |
| Diabetes | 2 | 3 |
| Hypertension | 4 | 5 |
| COPD | 0 | 1 |
| Coronary artery disease | 2 | 3 |
| Chronic kidney disease | 0 | 1 |
| Atrial fibrillation | 2 | 1 |
| OSA | 3 | 4 |
| $H_2$FpEF score | 5.4±1.9 | 5.6±1.8 |
| **Echocardiogram:** | | |
| LVEF (%) | 60.2±4.8 | 63.3±5.7 |
| Average E/E' | 10.9±3.2 | 11.6±1.3 |
| TAPSE (mm) | 18.8±3.7 | 16.9±4.2 |
| TAPSE/PASP (mm/mmHg) | 0.65±0.11 | 0.55±0.11 |
| **Rest Hemodynamics:** | | |
| mPAP (mmHg) | 28.6±4.3 | 30.8±7.6 |
| PAWP (mmHg) | 18.4±3.9 | 18.4±4.2 |
| Cardiac output (L/min) | 5.65±0.69 | 4.89±0.78 |
| PVR (Woods unit) | 1.8±0.3 | 3.5±0.8 |
| PAC (mL/mmHg) | 4.0±0.9 | 2.7±0.7 |
| **Exercise Hemodynamics:** | | |
| mPAP/CO slope | 3.5±1.2 | 7.2±4.1 |
| PAWP/CO slope | 2.3±0.2 | 4.1±2.0 |
| **Cardiopulmonary exercise test:** | | |
| Peak watts | 92.0±18.6 | 38.0±17.9 |
| Peak $VO_2$ (mL/Kg/min) | 13.6±3.6 | 9.4±2.9 |
| $V_E/VCO_2$ slope | 33.5±1.7 | 36.5±3.6 |
| OUES (mL/min/Log[L/min]) | 1.51±0.41 | 1.28±0.22 |
| **Cardiac MRI:** | | |

| | | |
|---|---|---|
| RVEF (%) | 51.8±7.5 | 42.2±9.5 |
| RV strain – longitudinal (%) | -20.2±2.7 | -19.6±0.9 |
| RV strain – circumferential (%) | -13.4±2.8 | -13.5±2.9 |
| LV strain – longitudinal (%) | -18.2±3.2 | -14.3±1.6 |
| LV strain – circumferential (%) | -19.5±1.0 | -16.4±2.7 |
| LA EF (%) | 37±10 | 26±4 |
| LA longitudinal strain (%) | 22.4±8.0 | 10.2±4.1 |

ABBREVIATIONS: PH-HFpEF: pulmonary hypertension due to heart failure with preserved ejection fraction, IpcPH: isolated post-capillary PH, CpcPH: combined pre-/post-capillary PH, BMI: body mass index, COPD: chronic obstructive pulmonary disease, OSA: obstructive sleep apnea, LVEF: left ventricular ejection fraction, Mitral E: mitral early diastolic velocity, E': mitral tissue doppler, TAPSE: tricuspid annular plane systolic excursion, PASP: pulmonary artery systolic pressure, mPAP: mean pulmonary artery pressure, PAWP: pulmonary artery wedge pressure, PVR: pulmonary vascular resistance, PAC: pulmonary artery compliance, CO: cardiac output, $VO_2$: oxygen consumption, $V_E/VCO_2$: ventilatory efficiency of carbon dioxide, OUES: oxygen uptake efficiency slope, RVEF: right ventricular ejection fraction, LV: left ventricular, LA: left atrium.

**Table S5. Sequencing and alignment metrics from PH-HFpEF risk groups.**

| Risk group | Total read count | Unaligned reads | polyA (%) | Mean read length (bp) |
|---|---|---|---|---|
| High-risk | 70.8 M | 18,615 | 98.5 | 1,550 |
| Low-risk | 68.3 M | 11,477 | 98.8 | 1,509 |

**Table S6. Titin transcript features**

† Exon 326 is 17106 bp— beyond the ≈10 kb HiFi insert length attainable with the PacBio Kinnex Iso-Seq workflow—so the putative splice junction (cell K39) at this locus could not be validated; its status is therefore reported as undetermined.
